# Small-cohort GWAS discovery with AI over massive functional genomics knowledge graph

**DOI:** 10.1101/2024.12.03.24318375

**Authors:** Kexin Huang, Tony Zeng, Soner Koc, Alexandra Pettet, Jingtian Zhou, Mika Jain, Dongbo Sun, Camilo Ruiz, Hongyu Ren, Laurence Howe, Tom G. Richardson, Adrian Cortes, Katie Aiello, Kim Branson, Andreas Pfenning, Jesse M. Engreitz, Martin Jinye Zhang, Jure Leskovec

**Affiliations:** Department of Computer Science, Stanford University School of Engineering, Stanford, CA, USA; Department of Genetics, Stanford University School of Medicine, Stanford, CA, USA; Basic Sciences and Engineering Initiative, Betty Irene Moore Children’s Heart Center, Lucile Packard Children’s Hospital, Stanford, CA, USA; Artificial Intelligence and Machine Learning, GSK, South San Francisco, CA, USA; Arc Institute, Palo Alto, CA, USA; Ray and Stephanie Lane Computational Biology Department, Carnegie Mellon University, Pittsburgh, PA, USA; Department of Human Genetics and Genomics, GSK, Stevenage, UK; Department of Human Genetics and Genomics GSK, Heidelberg, DE; The Novo Nordisk Foundation Center for Genomic Mechanisms of Disease, Broad Institute of MIT and Harvard, Cambridge, MA, USA; Gene Regulation Observatory, Broad Institute of MIT and Harvard, Cambridge, MA, USA; Stanford Cardiovascular Institute, Stanford University School of Medicine, Stanford, CA, USA

## Abstract

Genome-wide association studies (GWASs) have identified tens of thousands of disease associated variants and provided critical insights into developing effective treatments. However, limited sample sizes have hindered the discovery of variants for uncommon and rare diseases. Here, we introduce KGWAS, a novel geometric deep learning method that leverages a massive functional knowledge graph across variants and genes to improve detection power in small-cohort GWASs significantly. KGWAS assesses the strength of a variant’s association to disease based on the aggregate GWAS evidence across molecular elements interacting with the variant within the knowledge graph. Comprehensive simulations and replication experiments showed that, for small sample sizes (*N*=1-10K), KGWAS identified up to 100% more statistically significant associations than state-of-the-art GWAS methods and achieved the same statistical power with up to 2.67× fewer samples. We applied KGWAS to 554 uncommon UK Biobank diseases (*N*_case_ <5K) and identified 183 more associations (46.9% improvement) than the original GWAS, where the gain further increases to 79.8% for 141 rare diseases (N_case_ <300). The KGWAS-only discoveries are supported by abundant functional evidence, such as rs2155219 (on 11q13) associated with ulcerative colitis potentially via regulating *LRRC32* expression in CD4+ regulatory T cells, and rs7312765 (on 12q12) associated with the rare disease myasthenia gravis potentially via regulating *PPHLN1* expression in neuron-related cell types. Furthermore, KGWAS consistently improves downstream analyses such as identifying disease-specific network links for interpreting GWAS variants, identifying disease-associated genes, and identifying disease-relevant cell populations. Overall, KGWAS is a flexible and powerful AI model that integrates growing functional genomics data to discover novel variants, genes, cells, and networks, especially valuable for small cohort diseases.

## 1 Introduction

Human genetics evidence plays a crucial role in treatment development^1, 2^, yet the limited understanding of disease genetics currently poses a significant obstacle to effective drug discovery^3, 4^. Genome-wide association studies (GWAS) perform statistical tests to identify significant associations (e.g., *P* < 5 × 10^−8^) between millions of genetic variants and a phenotype (such as a disease or trait) across individuals in large populations, and this approach has uncovered tens of thousands of disease-associated variants and genomic regions (commonly called ‘loci’)^5, 6^. Subsequent functional studies of these findings have provided crucial insights into disease mechanisms and etiology ^1, 2, 5, 7–9^. Despite the potential of GWAS, their effectiveness in making novel discoveries about the genetic nature of diseases has been hindered by limited sample sizes, as GWAS requires large sample sizes that provide enough statistical power to identify causal genetic variants^5^. This requirement becomes a significant impediment when dealing with the many uncommon and rare diseases with lower frequencies, where large cohorts are challenging to assemble. Therefore, it is crucial to develop a more data-efficient approach to GWAS that maintains the same statistical power while requiring much smaller sample sizes. Such an approach would have a significant impact by enabling us to uncover the mechanisms of uncommon diseases reliably.

The rapid advancement of experimental techniques and functional genomics data offers new opportunities to address these limitations. These data have enabled the measurement of diverse molecular features in various cellular contexts^10–18^, such as chromatin accessibility, gene expression levels, and protein-protein interactions, providing an increasingly clear picture of cellular and molecular functions of a variant. Previous studies have integrated functional genomics data to improve GWAS association testing^19–25^ and downstream analyses^26–30^. However, the underlying methods^19–23^ primarily focus on a limited set of variant-level functional annotations^24, 25, 31, 32^, which may lack sufficient power for GWAS in small cohorts. Harnessing broader knowledge beyond variant-level annotations holds great potential to significantly enhance GWAS discovery and yield new biological insights.

Here, we present Knowledge Graph GWAS (KGWAS), a geometric deep learning method that integrates GWAS summary association statistics with massive functional genomics data to enable powerful association testing. KGWAS uses a functional genomics knowledge graph (KG) to capture relationships between genetic variants, aggregating diverse biological evidence to improve statistical power in distinguishing true disease-associated variants from spurious ones. KGWAS constructs a comprehensive KG across variants, genes, and gene programs to encode functional genomics knowledge, incorporating not only 70 variant annotations^19, 31^ but also 40,546 gene-level annotations^33^ and 11 million interactions from 55 relation types between variants, genes, and gene programs^12, 15, 17, 18, 34–40^. For a given disease, KGWAS trains a disease-specific graph neural network to predict disease associations for each variant based on the KG, and uses the predictions as a prior to refine association statistics through a p-value weighting framework^41^.

We showed via extensive simulations and replication experiments using the UK Biobank^42^ and FinnGen^43^ cohorts that KGWAS is well-calibrated and substantially more powerful than existing approaches. We applied KGWAS to GWAS summary statistics of 729 diseases and complex traits, including 413 uncommon diseases (300-5,000 cases) and 141 rare diseases (<300 cases). KGWAS identified 46.9% more statistically significant variants than the original GWAS across the 554 uncommon and rare diseases, and the gain further increased to 79.8% when restricting to the 141 rare diseases. We further leveraged KGWAS to systematically improve downstream applications, including using KGWAS-inferred disease-specific links to generate functional interpretations for disease variants and using KGWAS summary statistics to improve identification of disease-associated genes and disease-relevant cell populations. We validated our findings using independent GWAS cohorts, orthogonal experimental data, and expert-curated databases. Lastly, we provide a user-friendly interface for scientists to explore and analyze KGWAS discoveries, available at kgwas.stanford.edu.

## 2 Results

### Overview of methods

KGWAS integrates GWAS summary statistics with comprehensive functional genomics data to improve the power of genetic association testing. This approach may be effective because genetic variants with functional roles are more likely to contribute to disease^19–30^. Here, genetic variants are mutations on the human genome, loci are genomic regions (e.g., 10-500 kb), and GWAS tests for significant associations between genetic variants and diseases/traits in large populations^5^. KGWAS constructs a massive knowledge graph (KG) that captures functional properties and interactions between variants, genes, and gene programs (predefined groups of genes with shared functions). For a given disease, KGWAS trains an AI model to predict the association strength for each variant based on the KG, using these predictions as a prior to improve GWAS power. KGWAS improves association power for any GWAS method^44–48^, and the resulting KGWAS summary statistics are compatible with summary statistics-based downstream analyses. Further details are provided below and in the Methods section.

The KGWAS KG encompasses 784,708 UK Biobank directly genotyped variants^42^, 19,346 protein-coding genes, and 44,085 gene programs as graph nodes^18^; variant nodes are represented using 70 baseline-LD variant annotations^19, 31^ and gene nodes are represented using 40,546 gene annotations from scRNA-seq datasets^33^ (Figure 1a, Supplementary Tables 1,2; Methods). In addition, the KGWAS KG encodes comprehensive molecular interactions between variants, genes, and gene programs as graph edges, including (1) 8,629,515 variant-to-gene (V2G) links representing 14 relation types, such as activity-by-contact (ABC) enhancer-gene links and expression quantitative trait loci (eQTL) links^12, 15, 34–37^, (2) 2,330,109 gene-to-gene (G2G) links representing 40 relation types, such as genetic or physical interactions^17, 38–40^, and (3) 116,610 gene-to-program (G2P) links from the Gene Ontology^18^. Together, our KG represents one of the most comprehensive genome-wide maps of variant-to-gene-to-program functions.

**Figure 1:**
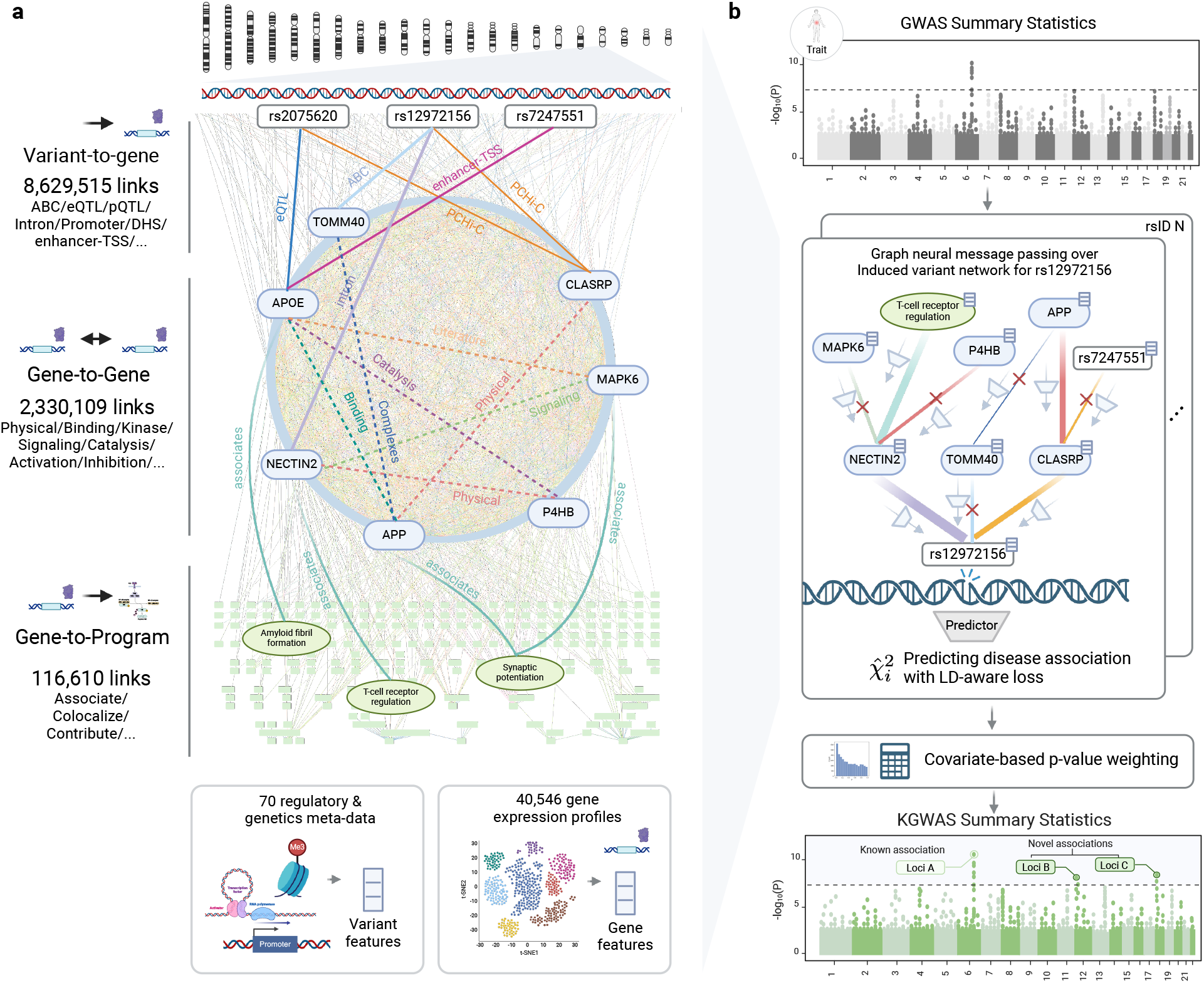
Overview of KGWAS. KGWAS is a deep learning framework that leverages a massive functional genomics knowledge graph to improve GWAS association testing and infer disease-critical networks. **a**. The KG nodes include 784,708 UK Biobank directly genotyped variants, 19,346 protein-coding genes, and 44,085 gene programs. Variant nodes are represented using 70 baseline-LD variant annotations and gene nodes are represented using 40,546 gene annotations from scRNA-seq datasets. The KG edges include 8,629,515 variant-to-gene links from 14 relation types, 2,330,109 gene-to-gene links from 40 relation types, and 116,610 gene-to-program links. KGWAS utilizes this large-scale KG to estimate the functional prior of each variant being causal for disease, effectively improving the accuracy of GWAS association testing. **b**. For a given disease, KGWAS takes GWAS summary statistics as input and outputs updated association statistics. It learns a heterogeneous graph attention neural network to aggregate functional evidence, estimating a variant’s prior relevance to disease and thereby improving association testing power. Specifically, for each variant (rs12972156 in this example), KGWAS trains the model to predict the variant’s marginal GWAS χ^2^ statistic using functional embeddings from its induced two-hop neighborhood network. LD is accounted for via an LD-aware loss function. Based on the trained network, KGWAS recomputes association p-values through a covariate-based p-value weighting approach, weighting the original GWAS p-values with predicted χ^2^ statistics that incorporate the variants’ aggregate functional evidence.

For a given disease, KGWAS integrates the KG with GWAS summary statistics to train a disease-specific graph neural network (GNN), producing more accurate association p-values (Figure 1b; Methods). First, KGWAS employs a heterogeneous GNN to encode variant, gene, and program nodes in the KG into biologically meaningful embeddings. Second, it learns disease-specific network weights through supervised learning to predict GWAS x^2^ association statistics based on variant embeddings, leveraging an LD-aware loss function to account for varying levels of LD (linkage disequilibrium) between variants^49^. Third, KGWAS produces a new set of association p-values by weighting the original GWAS p-values with predicted x^2^ statistics that reflect the variants’ aggregate functional evidence, leveraging a p-value weighting framework to ensure calibration^19, 41, 50^. We consider three downstream applications: (1) interpreting prioritized variants via the KGWAS-learned local disease network, (2) integrating KGWAS summary statistics with MAGMA^51^ to prioritize disease-associated genes, and (3) integrating KGWAS summary statistics with scDRS^52^ to detect disease-associated cell populations. We provide a human-centered graphical user interface to allow scientists to explore the discoveries made by KGWAS (kgwas.stanford.edu).

### Simulations assessing calibration and power

We evaluated the calibration and power of KGWAS using extensive null and causal simulations, comparing it to the non-functionally informed original GWAS and the state-of-the-art functionally-informed GWAS method FINDOR^19^ (Methods). We considered 542,758 UK Biobank variants that passed quality control^42^ and primarily focused on small-cohort GWAS, with sample sizes below 10K. We employed realistic genetic architectures. In our primary simulations, we considered *N* = 5,000 individuals, 20,000 causal variants, and heritability 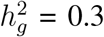; other settings were also evaluated. In null simulations, we placed all causal variants on odd chromosomes and used even chromosomes to evaluate calibration, similar to previous work^19^. In causal simulations, we selected 15 causal gene programs, randomly selected 100 genes connected to these programs as causal genes, and randomly selected 20,000 variants connected to these causal genes and programs as causal variants; to simulate realistic genetic architectures, we set the probability that a variant is causal to be proportional to the heritability enrichment of the variant’s functional annotations, as estimated from real data^31^. We computed the original GWAS association statistics using FastGWA^44^ when sample sizes >3K and switched to PLINK^45^ otherwise, following the FastGWA guideline^44^. We used the genome-wide significance threshold (*P* =5 × 10^−8^), and considered independent associations by pruning variants with LD *r*^2^ > 0.01 or within 0.1 centimorgan^53^. Further details are provided in the Methods section.

We reached 3 main conclusions. First, KGWAS was well-calibrated in null simulations, producing a similar level of false positives as the original GWAS and FINDOR (Figure 2a, Supplementary Table 3). Second, KGWAS detected substantially more independent true associations than the original GWAS and FINDOR in causal simulations (48.6% and 9.6% more discoveries, resp., Figure 2a, Supplementary Table 4). Third, we conducted additional simulations with alternative values of heritability (0.1, 0.2, and 0.5, instead of 0.3) and alternative numbers of causal variants (2K and 10K, instead of 20K) for both null and causal simulations, and determined that KGWAS remained well-calibrated and demonstrated consistently improved power (Supplementary Figures 1,2).

**Figure 2:**
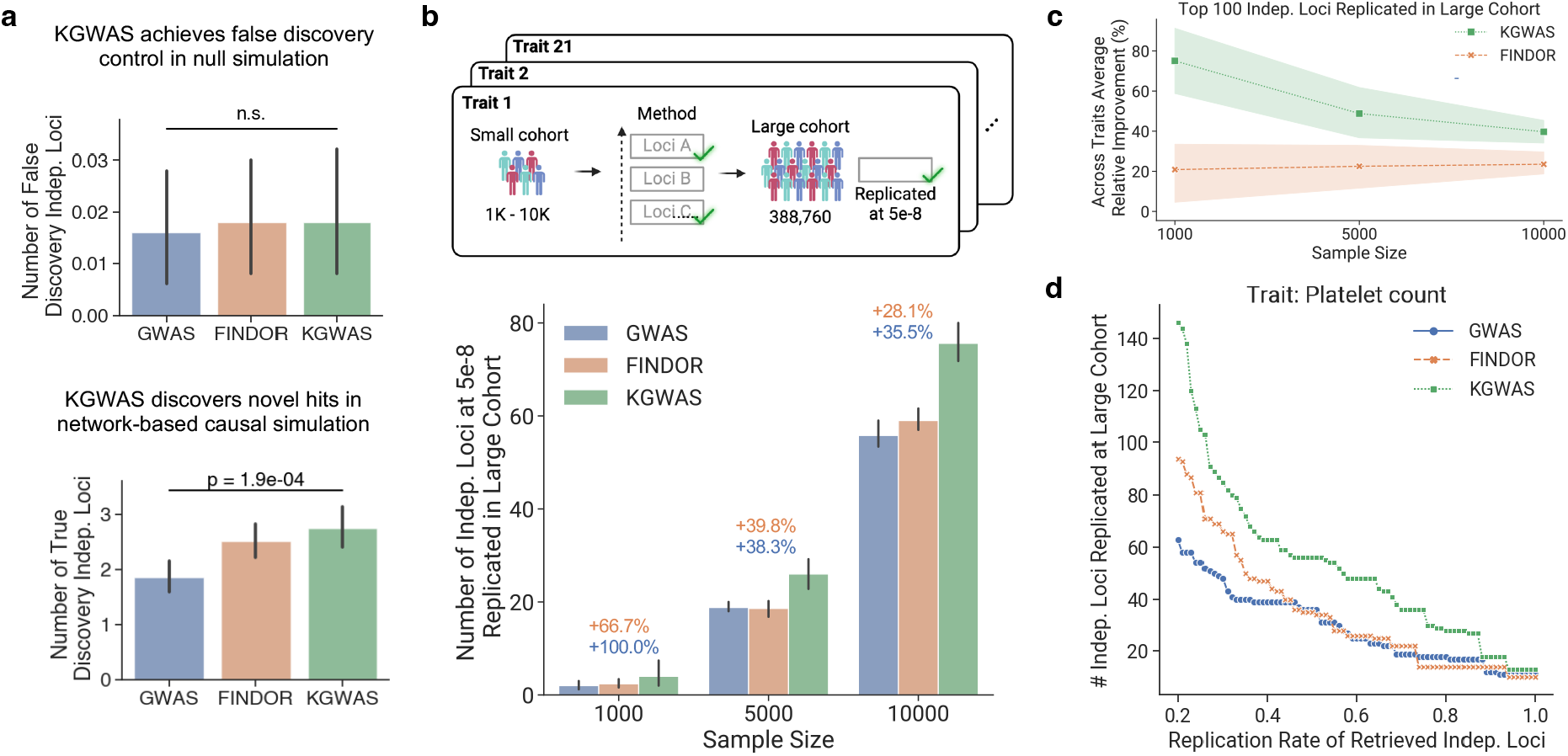
Results for simulations and replication experiments. **a**. Results for null and causal simulations.The upper panel shows results for null simulations, where the y-axis represents the number of false discoveries. KGWAS detected a similar number as baseline methods. The lower panel shows results for causal simulations, where the y-axis shows the number of true discoveries. KGWAS detected significantly more true associations than baseline methods. In both panels, error bars denote 95% confidence intervals around the mean of 500 simulation replicates. Numerical results are reported in Supplementary Tables 3,4. **b**. Results for replication experiments for genome-wide signficant loci (*P <*5 × 10^−8^). KGWAS identified more independent loci that were replicable in large cohorts compared to the original GWAS and FINDOR across different sample sizes. The x-axis denotes different sample sizes and the y-axis denotes the number of replicated discoveries averaged across 21 independent UK Biobank traits. Error bars denote 95% intervals around the mean of 5 replicates of the subsampled cohort. Numerical results are reported in Supplementary Table 6. **c**. Results for replication experiments for top 100 loci. The top 100 independent loci discovered by KGWAS had significantly higher replication rates across different sample sizes. The x-axis denotes different sample sizes and the y-axis denotes the number of replicated discoveries averaged across 21 independent UK Biobank traits. Error bars denote 95% intervals around the mean of 5 replicates of the subsampled cohort. Numerical results are reported in Supplementary Table 7. **d**. Replication rate at different thresholds for the trait platelet count. x-axis represents the proportion of independent loci replicated in the larger cohorts and y-axis represents the number of replicated loci in the larger cohorts. Additional results for simulations and replication experiments are reported in Supplementary Figures 1-6.

We conclude that KGWAS is well-calibrated and substantially more powerful than the original GWAS and FINDOR in realistic null and causal simulations.

### Systematic replication experiments across 21 diseases and traits

We conducted systematic replication experiments across 21 well-powered independent UK Biobank diseases/traits to provide more realistic evaluations (z-score >6 for nonzero SNP-heritability^37^; genetic correlation 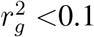^49^; average *N*=374K; Supplementary Table 5). We applied GWAS methods to subsampled cohorts (*N*=1K/5K/10K, typical for small-cohort GWAS) and assessed the replication on the full cohort (*N*=374K). We used FastGWA^44^ when sample sizes >3K and PLINK^45^ otherwise to conduct the original GWAS and considered independent associations, similar to the previous section. Further details are provided in the Methods section.

We reached 3 main conclusions. First, we considered the number of genome-wide significant loci (*P <*5 × 10^−8^) of each method that was replicated in the full cohort (original GWAS *P <*5×10^−8^). KGWAS produced substantially more replicated discoveries than the original GWAS (100.0%/38.3%/35.5% more replications at 1K/5K/10K resp.) and FINDOR (66.7%/39.8%/28.1% more replications at 1K/5K/10K resp.) (Figure 2b, Supplementary Table 6); the improvement was more pronounced at smaller sample sizes and consistent across diseases/traits (Supplementary Figure 3). Notably, KGWAS requires 1.24-2.67× fewer samples to achieve the same power as the original GWAS (375/3,953/8,035 vs. 1K/5K/10K; Supplementary Figure 4), highlighting its higher data efficiency. Second, we considered the top 100 loci of each method (instead of *P <*5 × 10^−8^ loci) that were replicated in the full cohort (original GWAS *P <*5 × 10^−8^). KGWAS again substantially outperformed both the original GWAS (75.0%/48.8%/39.7% more replications at 1K/5K/10K resp.) and FINDOR (50.4%/27.4%/15.5% more replications at 1K/5K/10K resp.) (Figure 2c, Supplementary Table 7), with the improvement being more pronounced at smaller sample sizes. Third, KGWAS consistently outperformed both the original GWAS and FINDOR across various significance thresholds (see Figure 2d for a representative example; average AUC improvements of 101.4%/68.1%/43.2% over the original GWAS and 77.6%/49.4%/8.2% over FINDOR at 1K/5K/10K resp.), suggesting the improvements were robust to different threshold choices.

We conducted 7 secondary analyses to rigorously assess KGWAS’s robustness and identify key performance drivers. First, we repeated the experiments with larger sample sizes (*N*=50K/100K /200K, instead of 1K/5K/10K; Supplementary Figure 5). KGWAS still outperformed the original GWAS and FINDOR, but the improvements were smaller (e.g., 13.3% more replications for *N*=200K vs. 27.7% for *N*=5K compared to the original GWAS), underscoring KGWAS’s strength in small-cohort GWAS. Second, we repeated the experiments for 15 traits in the independent FinnGen cohort ^43, 48^. KGWAS showed similar improvements over the original GWAS (180%/63% more replications for 5K/10K) and FINDOR (133%/56% more replications for 5K/10K) (no discoveries for 1K for all 3 methods; Supplementary Figure 6). Third, substituting the KGWAS KG with a randomized one resulted in worse performance which was similar to that of the original GWAS, confirming the KG’s role in driving performance gains (Supplementary Figure 7). Fourth, KGWAS’s performance progressively degraded as increasingly large random fractions of the KG were removed, underscoring the need for comprehensive modeling of molecular interactions (Supplementary Figure 8). Fifth, using alternative variant and gene embeddings from foundation models (e.g., ESM^54^, Enformer^55^) resulted in worse performance (Supplementary Figure 9), justifying our choices of variant and gene embeddings. Sixth, using pre-training objectives to generate variant embeddings resulted in worse performance, reinforcing the validity of our choice of disease-specific modeling (Supplementary Figure 10). Seventh, we determined that variants significant in KGWAS but not in the original GWAS had borderline association p-values in the original GWAS (from 1.44 × 10^−6^ to 5.01 × 10^−8^; Supplementary Figure 11), suggesting that KGWAS is effective at prioritizing likely replicable variants among those with borderline associations.

We conclude that KGWAS is substantially more powerful than both the original GWAS and FINDOR in replication experiments, with particularly notable improvements in small-cohort GWASs. The enhancement is robust and consistent across various thresholds, sample sizes, and datasets, demonstrating the benefit of integrating a massive functional genomics knowledge graph with summary statistics.

### Systematic discovery across 554 uncommon diseases in the UK Biobank

To expand genetic insights across uncommon and rare diseases comprehensively, we applied KGWAS to 554 diseases with fewer than 5K cases in the UK Biobank^42^ (Supplementary Table 8; Data availability). These diseases span 16 areas and include 141 rare diseases with fewer than 300 cases (Figure 3a). We used FastGWA^44^ for the original GWAS.

**Figure 3:**
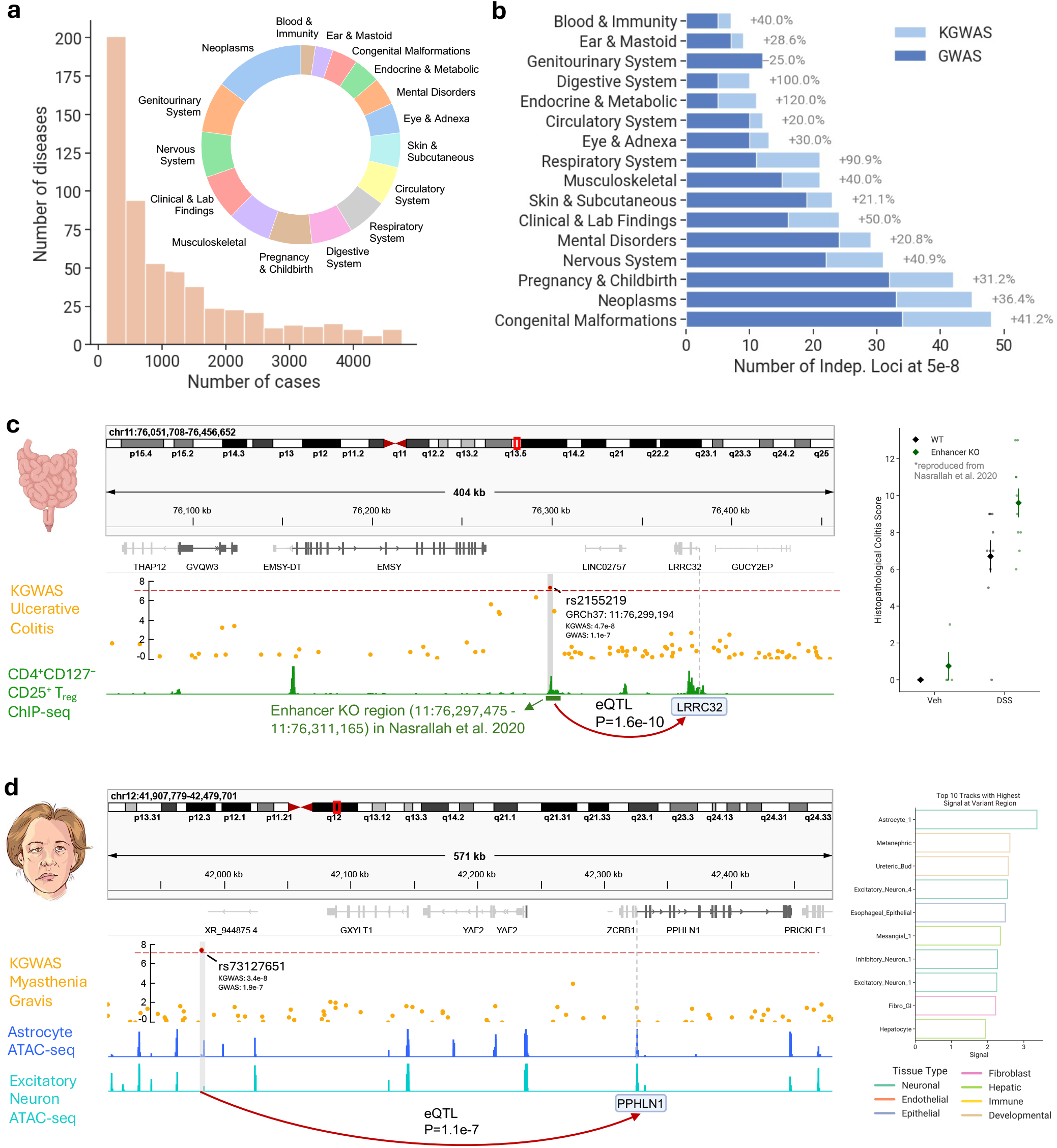
KGWAS discovers novel loci across 554 uncommon and rare diseases in the UK BioBank. **a**. Overview of 554 diseases. The x-axis is the number of cases and the y-axis is the number of diseases. The pie chart describes the proportion of diseases across disease areas. **b**. Comparison of novel loci discovered by KGWAS and the original GWAS across various disease areas. KGWAS identified more novel loci than the original GWAS. The improvement is consistent across disease areas. The x-axis represents the number of independent loci at *P* =5 × 10^−8^ and the y-axis represents different disease areas. **c**. Functional evidence for the association between the rs2155219 locus and ulcerative colitis (UC) (KGWAS *P* =4.7 × 10^−8^, original GWAS *P* =1.1 × 10^−7^). The left panel shows that this locus contains a distal enhancer specifically regulating the expression of *Lrrc32* in murine CD4^+^ regulatory T cells (Treg) and is protective against induced colitis. In the KGWAS KG, rs2155219 is uniquely linked to the gene *LRRC32* in the region through an eQTL. The right panel shows that knocking out the aligned enhancer region in mice induces macroscopic and histopathological features of colitis, especially in Treg-affected environment. The x-axis denotes the condition (Veh for vehicle and DSS for Dextran Sulfate Sodium), while the y-axis represents the histopathological scores of colitis. This panel is reproduced from previous work^62^. Error bars denote standard errors. **d**. Functional evidence for the association between the rs73127651 locus association and myasthenia gravis (MG) (KGWAS *P* =3.4 × 10^−8^), orignal GWAS *P* =1.9 × 10^−7^). The left panel shows that this locus is linked to the *PPHLN1* gene via an eQTL and shows enriched chromatin accessibility in neuron-related cell types. The right panel shows the top 10 cell types enriched for chromatin accessibility at this locus, based on a chromatin accessibility atlas of 222 primary human cell types. The x-axis represents normalized z-scores of the peak signal and the y-axis represents the cell types. Numerical results are reported in Supplementary Table 9.

KGWAS discovered 576 independent loci across 554 diseases (*P* <5 × 10^−8^), including 324 loci also significant in the original GWAS and 252 novel loci, representing a 46.9% increase in power compared to the 392 loci significant in the original GWAS (Figure 3b). The improvement was relatively uniform across categories, with the greatest increases observed for endocrine & metabolic disorders (120.0%), digestive system (100.0%), and respiratory system (90.9%). Notably, KGWAS uncovered at least one genetic discovery for 92 diseases that had no findings in the original GWAS, providing valuable insights for these challenging cases. For the 141 rare diseases, KGWAS identified 79.8% more genetic loci than the original GWAS (214 vs. 119), underscoring its potential to uncover discoveries in rare disease genetics. We leveraged discoveries in the GWAS Catalog ^56^ to validate our findings, focusing on 19 diseases with at least 1 overlap between GWAS Catalog loci and discoveries from either KGWAS or the original GWAS; a discovery is considered replicated if it is found in the GWAS Catalog. Among these 19 diseases, there were 73 discoveries made by both KGWAS and the original GWAS, 9 discoveries unique to KGWAS, and 3 discoveries unique to the original GWAS. 3 of 9 KGWAS-unique discoveries (*P*=3.7 × 10^−9^ for significant overlap, Fisher’s exact test) and 0 of 3 GWAS-only discoveries (non-significant) were replicated in the GWAS Catalog, suggesting that KGWAS-unique discoveries may be biologically meaningful. In summary, KGWAS identified a substantial number of discoveries across 554 uncommon diseases that were missed by the original GWAS and are potentially biologically meaningful.

### KGWAS reveals functionally-supported disease loci missed by the original GWAS

We next discuss representative examples of associations across the 554 uncommon UK Biobank diseases made by KGWAS but missed by the original GWAS. We focus on an uncommon disease, ulcerative colitis (UC), and a rare disease, myasthenia gravis (MG), chosen due to their critical unmet therapeutic needs. Two additional examples are provided in Supplementary Figure 12: the association between rs2070729 and Crohn’s disease (KGWAS *P*=2.0 × 10^−8^, original GWAS *P*=2.4 × 10^−7^) and the association between rs200482 and Sarcoidosis (KGWAS *P*=2.2 × 10^−8^, original GWAS *P*=2.2 × 10^−7^).

UC affects approximately 5 million people worldwide and currently lacks effective treatments^57, 58^. KGWAS discovered an association between rs2155219 (on 11q13) and UC (*P*=4.7 × 10^−8^), which was not significant in the original GWAS (*P*=1.1×10^−7^; Figure 3c); this locus was also discovered in a larger UC GWAS (16,315 cases, compared to 4,333 cases in the UK Biobank)^59^. This discovery is supported by multiple lines of functional evidence. First, rs2155219 resides in a locus associated with several other immune-related diseases, including Crohn’s disease and Psoriasis^59–61^. Second, the rs2155219 locus contains a distal enhancer specifically regulating the expression of *Lrrc32* in murine CD4+ regulatory T cells (T_reg_) and is protective against induced colitis^62^. Variants in this locus are associated with differential H3K27ac acetylation in CD4^+^ CD127^-^ CD25^+^ T_reg_ cells^63^, and knockout of the enhancer region in mice induces macroscopic and histopathological features of colitis^62^ (reproduced in Figure 3c). Consistent with this, in the KGWAS KG, rs2155219 is uniquely linked to *LRRC32* through a blood eQTL from eQTLGen^64^ (*P*=1.6e-10).

MG is a rare autoimmune disorder that affects approximately 20 out of every 100,000 individuals worldwide, and currently, there is no effective cure available^65^. KGWAS identified an association between rs73127651 (on 12q12) and MG (*P*=3.4 × 10^−8^), which was not significant in the original GWAS (*P*=1.9 × 10^−7^) and had not been discovered previously in the GWAS Catalog (Figure 3d). This discovery is supported by multiple lines of functional evidence. First, the rs73127651 locus showed strong chromatin accessibility in neuron-related cell types, including astrocytes, excitatory neurons, and inhibitory neurons in a single-cell chromatin accessibility atlas^66^ (1st, 4th, and 7th strongest signals among 222 primary human cell types in the dataset; Supplementary Table 9). These neuron-related cell types are potentially relevant to MG, as the disease primarily affects neuromuscular junctions^67^. Second, in the KGWAS KG, rs73127651 is uniquely linked to the gene *PPHLN1* through a blood eQTL in eQTLGen^64^ (*P*=1.1 × 10^−7^) and its locus contains an additional brain eQTL for *PPHLN1* (*P*=6.7 × 10^−3^)^68^ *PPHLN1* is involved in epigenetic silencing by the HUSH complex ^69^ and is ubiquitously expresesd in brain-related cell types^70^. Defects in HUSH complex function cause neuropathies that, like MG, are characterized by progressive muscle weakness^71^. Such defects cause the production of type I interferons, a signature associated with MG ^72^. Currently, the mechanisms underlying MG are poorly understood^73^. Our findings suggest that the rs73127651 locus may harbor a causal variant (either rs73127651 or a variant in LD with it) that contributes to MG by regulating the expression of *PPHLN1* in neuron-related cell types, shedding light on the etiology of this rare disease.

In summary, KGWAS has identified many disease-associated loci that were missed by the original GWAS. These novel findings are supported by functional evidence, making them likely to be biologically meaningful.

### Evaluating accuracy of KGWAS disease-specific link prioritization

In addition to association testing, KGWAS further prioritizes disease-specific links for each variant using an attention-based graph interpretability method, providing critical mechanistic insights. We evaluate the accuracy of KGWAS-prioritized disease-specific variant-to-gene (V2G), gene-to-gene (G2G), and gene-to-program (G2P) links using simulations and real-world data, comparing it with the state-of-the-art graph XAI method GSAT^74^ and a uniform baseline that randomly selects links in the KG (Methods).

We first assessed KGWAS’s ability to prioritize disease-specific causal links using causal simulations in the subsection “Simulations assessing calibration and power”. We reached 3 main conclusions. First, at large sample sizes (*N*=374K), KGWAS was significantly more accurate than baseline methods in prioritizing causal links (including V2G, G2G, G2P) (AUPRC of 0.269, 0.069, 0.006 for KGWAS, GSAT, and the uniform baseline, resp.; Figure 4a, Supplementary Table 10). Moreover, KGWAS consistently assigned higher network weights to truly causal links than to non-causal links (64.3%, 96.7%, and 216.2% higher for V2G, G2G, and G2P links, resp.; Supplementary Figure 13). Second, at smaller sample sizes (*N* =10K,5K), KGWAS had slightly worse performance but continued to outperform baseline methods (KGWAS AUPRC of 0.264 for 10K and 0.151 for 5K; Supplementary Figure 14), indicating that KGWAS can provide meaningful variant interpretation at low sample sizes. Third, we confirmed that KGWAS ‘s causal link prioritization was the most robust compared to alternative strategies (details in Supplementary Figure 15).

**Figure 4:**
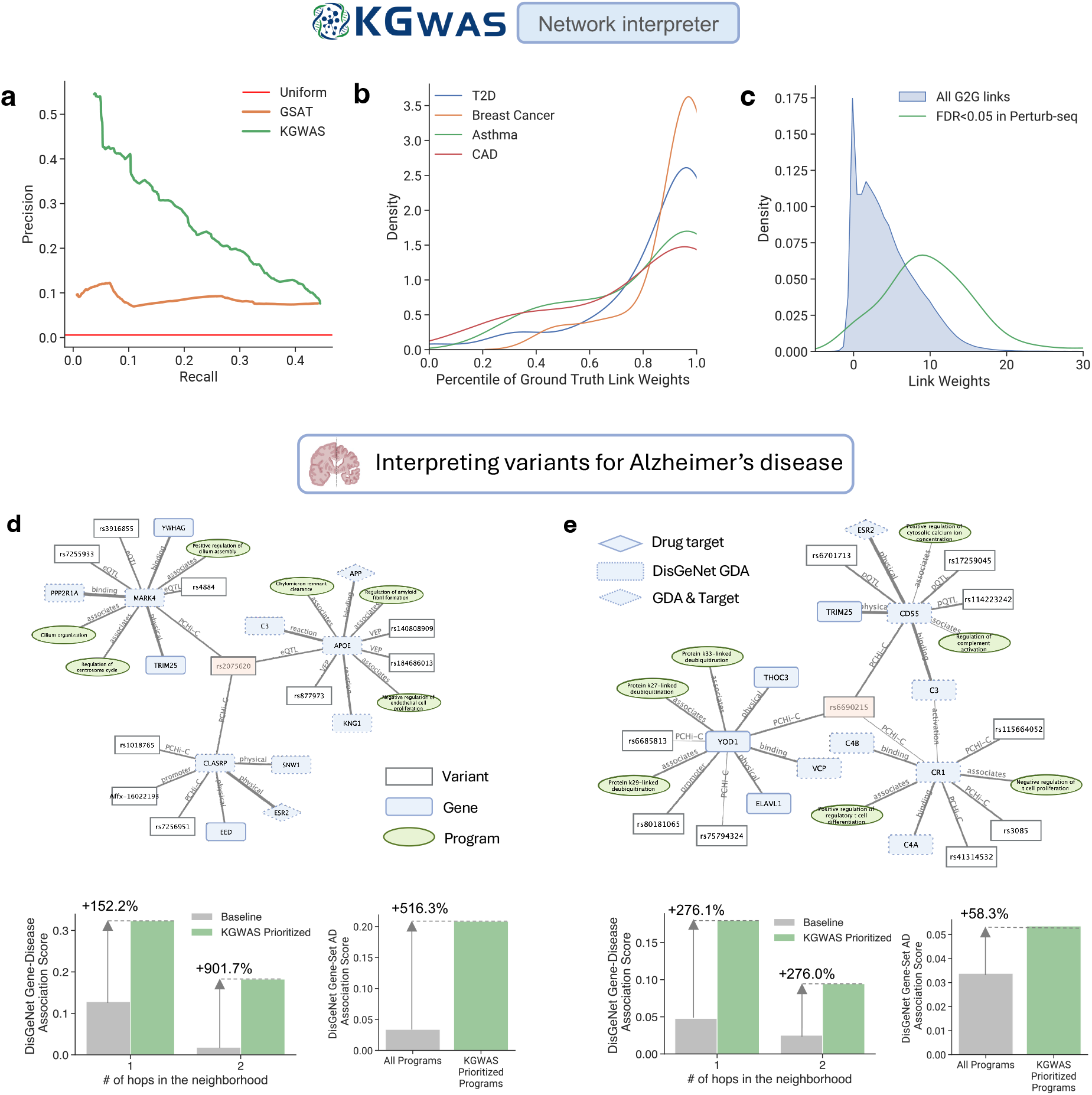
Results for KGWAS disease-critical link prioritization. **a**. Evaluation of disease link prioritization in causal simulations. KGWAS outperformed GSAT and the uniform baseline in identifying truly causal links. Numerical results are reported in Supplementary Table 10. **b**. Distribution of KGWAS weights of ground truth links across 4 diseases (T2D, breast cancer, asthma, CAD). Numerical results are reported in Supplementary Table 11. KGWAS consistently assigned high weights to gold-standard V2G links for these diseases. The y-axis represents the density, and the x-axis shows the percentile ranking of the ground truth V2G links. **c**. Distribution of KGWAS weights for G2G links. Perturb-seq G2G links (used as ground-truth) have higher KGWAS link weights compared to all G2G links of the same trait. The x-axis represents KGWAS link weights, and the y-axis represents the density. Numerical results are reported in Supplementary Table 12. **d-e**. KGWAS interpretation for two Alzheimer’s disease (AD) loci rs2075620 (panel d) and rs6690215 (panel e), both identified by KGWAS but missed by the original GWAS. Top panels visualize the KGWAS prioritized local disease networks. The bottom panels show the enrichment of KGWAS-prioritized genes and programs compared to AD-related genes and programs in DisGeNet (considered as gold-standard). The x-axis represents different sets of KGWAS-prioritized genes and programs, and the y-axis represents the enrichments compared to the corresponding genes and programs in DisGeNet. Numerical results are reported in Supplementary Table 13,14.

We next evaluated KGWAS-inferred disease-specific links using orthogonal datasets, focusing on 5 UK Biobank disease/traits where such data was available: type II diabetes (T2D; *N*_case_=30K), breast cancer (*N*_case_=14K), asthma (*N*_case_=37K), and coronary artery disease (CAD; *N*_case_=40K), and mean corpuscular hemoglobin (MCH; continuous trait; *N*=396K). First, we compared the KGWAS-inferred V2G links with expert-curated links from Open Targets^75^ for T2D, breast cancer, asthma, and CAD (Figure 4b and Supplementary Table 11). The Open Targets links were strongly enriched among KGWAS prioritized links (*OR* =11.2, 17.3, 8.0, 6.0 and *P*=1.5×10^−12^, 2.5×10^−7^, 3.1×10^−3^, 7.9×10^−4^ for T2D, breast cancer, asthma, and CAD, resp.),suggesting that KGWAS was effective at prioritizing likely causal V2G links. Second, we validated the KGWAS-inferred G2G links for MCH using a Perturb-seq dataset on a matched hematopoietic cell line^76^, where Perturb-seq G2G links were defined as links between the perturbed genes and the corresponding downstream genes (Figure 4c and Supplementary Table 12). KGWAS prioritized G2G links had strong overlap with the Perturb-seq links (*OR*=7.4, *P*=7.1 × 10^−128^), suggesting that KGWAS was effective at identifying causal G2G links.

We conclude that KGWAS accurately prioritizes relevant links for disease-associate variants among links in the KG, significantly outperforming alternative methods such as GSAT.

### KGWAS reveals molecular mechanisms for Alzheimer’s disease loci

We leverage KGWAS disease-specific link prioritization to provide mechanistic interpretations for disease-associated variants. We applied KGWAS to one of the largest Alzheimer’s disease (AD) GWAS summary statistics^77^ (*N*_case_=54,162, *N*_ctrl_=1,136,071, 135,479 variants shared with KGWAS KG), selected for its significant clinical relevance and widespread interest in medical research^78^. KGWAS discovered 19 independent loci, 11.7% more than the original GWAS (17 loci). Additional examples are provided in Supplementary Figures 16,17 for asthma and CAD. We next discuss two notable examples.

First, KGWAS detected a significant association between rs2075620 (on 19q13) and AD (*P*=4.6 × 10^−8^), which was not significant in the original GWAS (*P*=8.0 × 10^−8^); the rs2075620 locus harbors previously-reported AD associations (19q13)^79^ (Figure 3d, Supplementary Figure 18, and Supplementary Table 13). Among rs2075620 links in the KGWAS KG, KGWAS prioritized links were enriched with AD-related genes from DisGeNet^80^ (152.2%/901.7% enrichment for genes that are at most one or two edges away, resp.). KGWAS-prioritized gene programs also showed a 516.3% higher enrichment of AD-related genes than other programs in the general network. Notably, KGWAS linked rs2075620 to *APOE* (via eQTL), a gene pivotal to AD^81^, and further to *APP* (via binding interactions with *APOE*), another known AD drug target involved in amyloid plaque formation^82^. Additionally, KGWAS connected rs2075620 to pathways crucial in AD, including chylomicron remnant clearance and amyloid fibril formation regulation, whose dysregulation can impact lipid metabolism and promote amyloid-beta accumulation to contribute to neurodegeneration^83, 84^.

Second, KGWAS detected a significant association between rs6690215 (on 1q32) and AD (*P*=2.0 × 10^−8^), which was not significant in the original GWAS (*P*=2.0 × 10^−8^); the rs6690215 locus harbors previously-reported AD associations (1q32)^85^ (Figure 3e, Supplementary Figure 18, and Supplementary Table 14). Among rs6690215 links in the KGWAS KG, KGWAS prioritized links were enriched with AD-related genes from DisGeNet^80^ (276.1%/276.0% enrichment for genes that are at most one or two edges away resp.). KGWAS-prioritized gene programs also showed a 58.3% higher enrichment of AD-related genes than other programs in the general network. Notably, KGWAS linked rs6690215 to *CD55* and *CR1* (via Promoter Capture Hi-C), both regulating the complement system, a key component of AD-related neuroinflammation^86^. *C3, C4A*, and *C4B* as well as pathways involved in complement activation and T-cell proliferation are also in the local disease network for this variant, reinforcing its connection to complement activity and neuroinflammation^87^. Together, these findings suggest an immune-related mechanism for this variant.

In summary, KGWAS detects substantially more AD loci than the original GWAS and prioritizes plausible molecular mechanisms for these loci.

### KGWAS improves accuracy for summary-statistics-based downstream analyses

The KGWAS summary statistics are compatible with summary-statistics-based GWAS downstream analyses and could offer significant performance gains. We evaluated this in two widely-used analyses based on GWAS summary statistics: identifying disease-associated genes with MAGMA^51^ and detecting disease-relevant cell populations with scDRS^52^ (Methods).

We conducted two analyses for MAGMA-based disease-gene prioritization. First, we applied MAGMA to both the KGWAS and original GWAS summary statistics for a small 1K cohort across 21 independent UK Biobank diseases/traits (Supplementary Table 5), assessing the replication of the top 1,000 prioritized genes based on discoveries in the full cohort (average *N*=374K, FWER<0.05, using the original GWAS). Results are reported in Figure 5a and Supplementary Table 15. The top 1,000 prioritized genes using KGWAS had a 28.3% higher replication rate than those using the original GWAS, indicating more accurate disease-gene identification. The performance gain is consistent across sample sizes, thresholds, and traits (Supplementary Figure 19). Second, we compared the top 1,000 prioritized genes per trait against gold-standard drug targets^88^ across a separate set of 15 diseases where such information was available (average *N*_case_=19K). The prioritized genes using KGWAS exhibited a 17.2% higher overlap with drug targets compared to those using the original GWAS, highlighting KGWAS’s potential to identify putative drug targets (Figure 5b, Supplementary Table 16).

**Figure 5:**
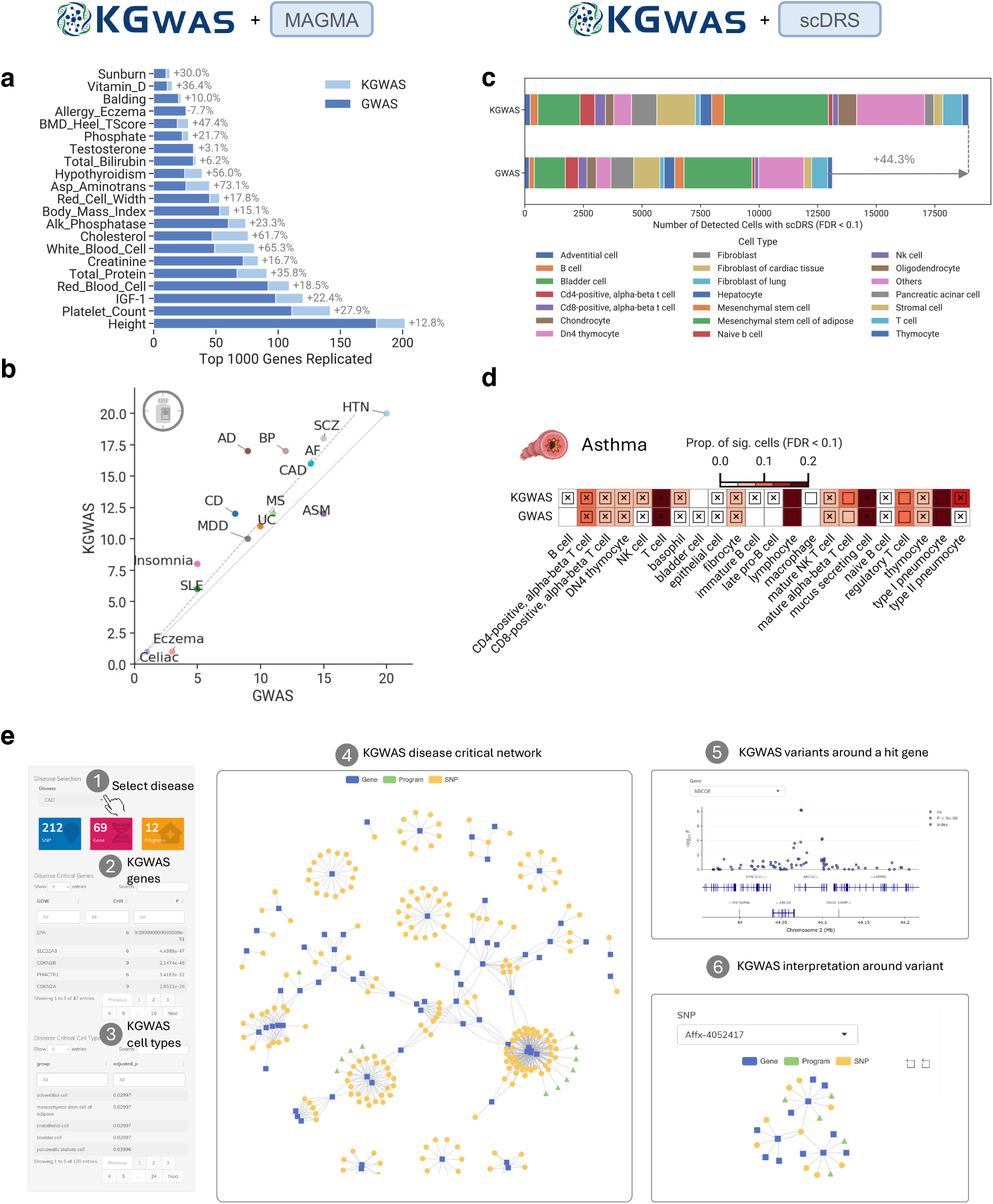
Results for using KGWAS to prioritize disease-critical genes and cell populations. **a**. Results for replication experiments on gene prioritization. KGWAS identifies more genes than the original GWAS that are replicable in large cohorts when integrated with MAGMA. The x-axis denotes the number of the top 1,000 genes that are replicated. Numerical results are reported in Supplementary Table 15. **b**. Results for validation with gold-standard drug target genes. KGWAS demonstrates more accurate prioritization of known drug target genes compared to GWAS. The y-axis and x-axis represent the number of drug-target genes for KGWAS and GWAS respectively. Numerical results are provided in Supplementary Table 16. **c**. Results for detection of disease-critical cell populations by integrating KGWAS with scDRS. KGWAS identifies more disease-critical cell populations on the Tabula Muris scRNA-seq atlas compared to the original GWAS. The x-axis is the number of significant disease-cell associations, colored by cell types. Numerical results are provided in Supplementary Table 18. **d**. Discovery of novel asthma-related B-cell clusters. KGWAS uncovers novel B-cell clusters (immature B cells and late pro-B cells) missed by the original GWAS. Numerical results are provided in Supplementary Table 18. **e**. A human-centered graphical user interface to facilitate the understanding of disease etiology. This interface allows scientists to explore KGWAS predictions including disease-critical variants, genes, cells, and networks. The interface is openly accessible at kgwas.stanford.edu.

We next assessed the performance of scDRS-based disease-cell identification, which, given GWAS summary statistics and scRNA-seq data, identifies cells that exhibit excess expression across a set of disease-associated genes informed by GWAS^52^. We analyzed 93 UK Biobank diseases (>10K cases or *P* <0.05 for non-zero SNP-heritability; Supplementary Table 17; Methods) in conjunction with the Tabula Muris Senis scRNA-seq dataset^89^ (110,824 cells from 120 cell types). Results are reported in Figure 5c,d and Supplementary Table 18. First, applying scDRS to KGWAS summary statistics identified 44.3% more disease-cell associations than that using the original GWAS, indicating a substantial increase in detection power (Figure 5c). Second, KGWAS-based analysis identified novel disease-cell associations missed by that using the original GWAS, such as a population of oligodendrocytes linked to nicotine use (Figure 5d). Oligodendrocytes, which myelinate neurons in the CNS, are increasingly recognized for their role in the cognitive effects of nicotine exposure^90^. In addition, KGWAS detected B cell populations (B cells, immature B cells, and late pro-B cells) associated with asthma, missed by the original GWAS. These cell populations are critical in asthma pathogenesis, and targeting of B cells and Immunoglobulin E, an antibody they produce, leads to the development of the medication omalizumab^91^ to treat severe allergic asthma unresponsive to standard therapies. This highlights KGWAS’s strength in uncovering disease-critical cell populations with therapeutic relevance.

In summary, the KGWAS summary statistics provide significant improvements in identifying disease-associated genes and cell populations.

### User-friendly interface enables rapid exploration of disease variants and mechanisms

We developed a user-friendly graphical user interface to facilitate hypothesis generation for loci discovered by KGWAS for 726 diseases in the UK Biobank (Figure 5e). For a given disease, scientists can explore an interactive mechanistic network that encompasses disease-specific genome-wide variant-to-gene-to-program links identified by KGWAS, facilitating the integration of functional evidence with disease variants, the discovery of gene modules, and an improved understanding of the genetic bases of diseases. Scientists can now perform variant interpretation for all 1,982 loci across 726 diseases in the UK Biobank by selecting a specific variant and visualizing the KGWAS-inferred disease-specific network associated with the given variant. Additionally, the interface enables scientists to find KGWAS disease-associated variants, genes, and cell types. This user-friendly interface is openly accessible at kgwas.stanford.edu.

## 3 Discussion

We have introduced KGWAS, a geometric deep learning method that enhances GWAS association testing and generates mechanistic hypotheses by reasoning over a comprehensive functional genomics knowledge graph. This KGWAS knowledge graph, one of the largest constructed to date, encompasses 11 million links between variants, genes, and gene programs. We showed via extensive simulations and replication experiments that KGWAS is well-calibrated and powerful in realistic scenarios. We applied KGWAS to 729 diseases/traits, including 403 uncommon diseases and 141 rare diseases, to detect novel disease-associated loci and uncover potential disease mechanisms. These findings provide valuable insights into the genetic basis of common and rare diseases and may inform the development of treatments. In addition, KGWAS consistently improves down-stream analyses based on GWAS summary statistics, such as identifying disease-associated genes and relevant cell populations. Integration of additional human genetics and functional genomics data using approaches such as KGWAS will continue to advance our understanding of disease genetics and facilitate follow-up studies.

We note several limitations and future directions of our work. First, KGWAS currently models only the 784,708 UK Biobank directly-genotyped variants. Expanding this set to include wholegenome variants could significantly improve the discovery and interpretation of disease-associated variants, particularly rare variants, which may have greater clinical importance. Second, the computational cost of KGWAS scales linearly with the number of variants and the number of edges per variant, making it highly scalable for analyzing a broader set of genetic variants and functional genomics data. Third, since the genotyped variants likely do not include the causal variants for disease, the KGWAS identified variants may tag the disease loci but may not represent causal variants themselves. Fourth, KGWAS updates p-values but does not provide signed effect sizes; extending the framework to estimate signed effect sizes, such as using updated signed z-scores^92^, would greatly improve its utility. Fifth, the KGWAS knowledge graph relies on functional genomics data such as eQTL links and gene ontology, which may be biased towards common and well-studied variants and genes. Sixth, the KGWAS knowledge graph is context-agnostic; expanding the framework to incorporate context-specific or cell type-specific networks would improve both discovery power and interpretability. Seventh, KGWAS does not yet provide p-values for its inferred disease-specific links; developing a rigorous statistical testing framework for these links will allow for more robust or trustworthy variant interpretation. Eighth, while we focus on the European populations, extending KGWAS to GWAS data from other populations could lead to new biological discoveries^93, 94^. Ninth, while we used KGWAS to prioritize genes and cell populations as as examples of downstream applications, the method can serve as a drop-in replacement for most analyses utilizing GWAS summary statistics. Despite these limitations, KGWAS was designed to be highly modular, allowing for easy integration of alternative entity embeddings, knowledge graphs, and variant sets. In summary, KGWAS is a powerful tool for identifying novel disease-critical variants, genes, cell populations, and networks, particularly in small-cohort GWAS.

## 4 Methods

### GWAS datasets and analyses

We considered individuals with European ancestry from the UK Biobank, analyzing the set of *N*=407,833 related individuals or the set of *N*=332,781 independent individuals. Specifically, following Bycroft et al.^42^, we restricted to individuals with self-reported White British ancestry (Field ID: 22006). For the set of independent individuals, we further performed pruning based on genetic relatedness (Field ID: 22018). Finally, we excluded individuals exhibiting sex chromosome aneuploidy (Field ID: 22019), individuals with mismatches between self-reported and genetically determined sex (Field IDs: 31 and 22001), individuals flagged for recommended genomic analysis exclusions due to signs of insufficient data quality (Field ID: 22010), and participants withdrawn from the study. We considered a set of 542,758 directly genotyped UK Biobank variants, filtered based on genotyping missingness lower than 0.02, Hardy-Weinberg equilibrium (HWE) p-value greater than 1 × 10^−8^, and minor allele frequency (MAF) greater than 0.01. Following Kichaev et al.^19^, we additionally excluded the MHC region on chromosome 6. By default, We used fastGWA^44^ for GWAS analysis when the sample size is greater than 3,000 and used PLINK^45^ otherwise, following the fastGWA guideline^44^. As covariates, we included the top 15 population PCs, sex, age, and assessment center status. For the linear model using PLINK, we removed additional related individuals based on KING relative scores^95^, selecting the first individual in each relative group. We used publicly available GWAS summary statistics for diseases analyzed in the subsection “KGWAS reveals molecular mechanisms for Alzheimer’s disease loci” and used individual-level data for all other analyses (Supplementary Table 19).

### Knowledge graph construction

Our knowledge graph aggregates information from 6 diverse databases to link variants to genes to programs. From variant-to-gene links, we integrate Open Targets Genetics^75^ and cS2G strategies^37^. For gene-to-gene links, we aggregate from BioGRID^17^, the Database of Interacting Proteins^40^, and STRING^38^. For gene-to-program links, we utilize gene ontology^18^. We standardize terms and merge edge types when necessary. All KG statistics can be found in Supplementary Tables 1,2.

### KGWAS problem formulation

Consider a GWAS study for a disease *d* on a set of variants 𝒮 with a summary statistics **p** ∈ [0, 1]^|𝒮|^. *p*_*i*_ is the p-value for SNP *i*, measuring association evidence to the disease. We are given a heterogeneous knowledge graph of variants, genes, and programs *G* = (𝒱, ε, 𝒯_*R*_) with nodes in the vertex set *v*_*i*_ ∈ 𝒱, edges *e*_*i,j,r*_ = (*v*_*i*_, *v*_*j*_, *r*) in the edge set ε. An edge is denoted by a triplet (*v*_*i*_, *v*_*j*_, *r*), where *r* ∈ 𝒯_*R*_ indicates the relation type, *v*_*i*_ is called the source node and *v*_*j*_ is called the target node. We model the relationship because each edge, such as a variant-to-gene edge, can be measured by multiple relationships, such as eQTL or ABC. Each node also has an initial embedding, which we denote as 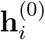. The goal is to learn a function 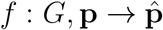 that takes in *G* and **p** and generates an adjusted p-values 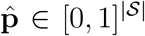. KGWAS consists of three modules: (1) heterogeneous graph neural network encoder; (2) prediction of tagged variance; (3) co-variate-based p-value weighting. After training, we also describe an interpretability procedure to obtain network importance scores. We describe each module in detail below.

### Heterogeneous graph neural network encoder

Given a knowledge graph, we aim to learn a numerical vector (i.e., network embedding) for each variant, gene, and program such that it captures biomedical knowledge encapsulated in the neighboring relational structures. This is achieved by transforming initial node embeddings through several layers of local graph-based non-linear function transformations to generate embeddings^96, 97^. These functions are optimized iteratively, given a loss function to propagate GWAS signals. Upon convergence, optimized functions generate an optimal set of node embeddings.

#### Step 1: Initialization

We denote the input node embedding **X**_*i*_ for each node *i*. We initialize the variant embedding with baseline-LD features (e.g. epigenetics, allele frequency, etc.) from Kichaev et al^19^. We initialize the gene embedding from the PoPS features^33^. Gene program embeddings are initialized randomly. Alternative embeddings are explored in Supplementary Figure 9. For every layer *l* of message-passing, there are the following three steps:

#### Step 2: Propagating relation-specific neural messages

For every relation type, KGWAS first calculates a transformation of the node embedding from the previous layer **h**^(*l-*1)^, where the first layer **h**^(0)^ = **X**, the initial embedding. This is achieved via applying a relation-specific weight matrix 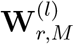 on the previous layer embedding:

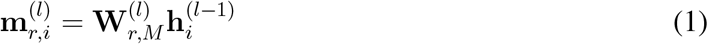

Then, based on this embedding, we can calculate the attention for each edge (*v*_*i*_, *v*_*j*_, *r*), which measures the importance of this edge for message-passing. The attention weight is based on the updated node embeddings from the source and target node, and is calculated via another relation-specific weight matrix 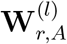:

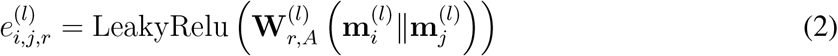

#### Step 3: Aggregating local network neighborhoods

For each node *v*_*i*_, we aggregate on the incoming messages 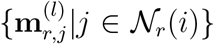 from neighboring nodes of each relation *r* denoted as 𝒩_*r*_(*i*). To leverage the edge attention, we want the aggregated node embedding for target node *v*_*i*_ to contain a higher contribution from neighborhood nodes that have high attention weight edges connected to it. Thus, first, we can apply a softmax function to normalize the attention weights that point to the target node:

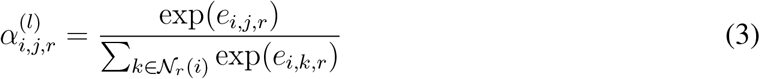

This *α* measures the edge-level contribution from the neighborhood and we can thus weight the importance of the node embedding messages based on these scores during aggregation:

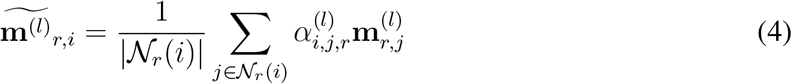

#### Step 4: Updating network embeddings

We then combine the node embedding from the last layer and the aggregated messages from all relations to obtain the new node embedding:

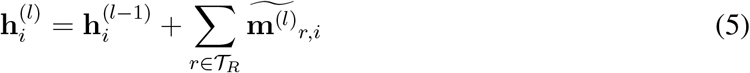

After *L* layers of propagation, we arrive at our encoded node embeddings **h**_*i*_ for each node *i*.

### LD-aware loss to predict tagged variance

The goal of this module is to propagate the GWAS signals over the KG so that the embedding reflects it if a variant/gene/program is enriched for disease association. The natural way to enable this is to update the variant embedding by predicting the GWAS signal. We use the χ^2^ statistics of a variant as the optimization target. χ^2^ statistics also correspond to the tagged phenotypic variance. Formally, given the GWAS summary statistics, we first applied a prediction head on the variant embedding 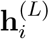 and obtained the predicted 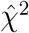. The predicted tagged variance 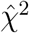 measures how much this variant tags the phenotype-enriched part of the knowledge graph. If it overlaps with many enriched genes, programs, and other variants in its local neighborhood, we have a stronger prior that this variant is enriched for signal.

As the χ^2^ statistics at SNPs in LD are correlated and the χ^2^ statistics of variants with high LD Scores have higher variance than the χ^2^ statistics of variants with low LD Scores, if we optimize by treating variants equally, it could bias the learning for SNPs in large LD block. To counterbalance it, we follow Bulik et al.^49^ and propose an LD-aware loss:

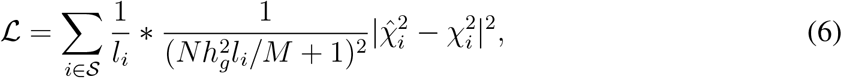

where *l*_*i*_ is the LD score for variant *i, M* is the number of variants in the reference LD panel, *N* is the sample size, and 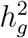 is the heritability constant. We then optimized the GNN weight 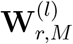based on this loss. In our analyses, we compute LD scores using in-sample LD when such information is available, such as for UK Biobank traits and use an ancestry-matched reference panel otherwise, such as using the UK Biobank LD score when analyzing publicly available summary statistics from the European population.

### Co-variate based p-value weighting and calibration

The predicted 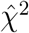 measures network priors but is hard to interpret for discovery since there is no false discovery control. In this module, we convert the prior into an adjusted p-value such that false discovery is controlled. We used the covariate-based p-value weighting framework^41^, following Kichaev et al^19^. Particularly, we stratify the SNP set 𝒮 into *B* even-sized bins based on the network prior 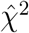. We use a null estimator^98^ to estimate the null proportion 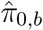 and alternate proportion 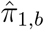 of each bin by fitting a cubic spline to p-value histogram. We then weight the p-value **p**_*b*_ in each bin *b* by a normalized weight^19^:

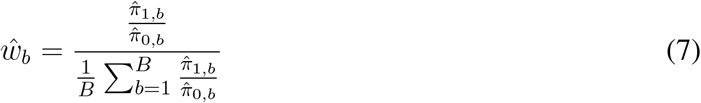

such that

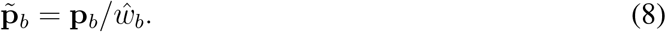

A bin with a larger number of alternate proportions will adjust the p-value to be more significant and vice versa. This procedure in theory preserves control of type 1 error with high probability^50^. Empirically, under simulation study, when the sample size is large, we observed strong control of false discovery. When the sample size is small (e.g., less than 10K), we observed a small inflation in false discovery. To ensure conservative false discovery control, we conducted an additional step of calibration with the original GWAS p-values. Particularly, we applied a rank-preserving scaling factor γ such that 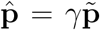 and the γ is selected such that the number of discoveries is the same with the original GWAS between a region [*p*_*a*_, *p*_*b*_]. *p*_*a*_, *p*_*b*_ is set to be a large p-value threshold so that we do not affect the true discovery number under a significant threshold. We selected *p*_*a*_ = 1*e −* 3, *p*_*b*_ = 1*e −* 2 using a set of 50 validation random states and found it to be robust when extending to larger 500 random states run and across other configurations.

### Network importance scores

To estimate disease critical network, we built upon the pre-softmax attention score α_*i,j,r*_ for edge between node *i, j* in relation type *r* as a basis. The score is high when it plays an important role when predicting χ^2^, thus, corresponding to a measure of association for the KG link to the phenotype. As our graph is heterogeneous across relation types and the attention score is computed based on per-relation weight, we observe differences in the distribution of attention scores across relation types. A relation type with a large mean can override another relation type with a small mean when doing ranking. To normalize it, we compute an updated network importance score as the relation-wise z-score: 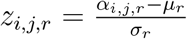, where *µ*_*r*_, σ_*r*_ is the weighted average and standard deviation for the relation *r*. As there could exist many relation types between two nodes, and we are interested in finding the most useful relation, we pick the highest score across relations to represent the edge importance 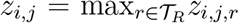. See Supplementary Figure 15 for benchmarks on other importance scores including GNN XAI methods and variations of weighting approaches.

### Details for null and causal simulations

We used GCTA^99^ to simulate continuous phenotypes under a polygenic model with normally distributed causal effect sizes and a specified number of causal variants and heritability. We used realistic estimates of these parameters (number of variants 2K, 10K, and 20K) and heritability (0.1, 0.2, 0.3, 0.5). We sampled *N*=5K individuals from the UK Biobank as the real genotypes for small-cohort GWAS. In null simulations, we placed all causal variants on odd chromosomes and computed the number of discoveries in the even chromosomes as the false discoveries to evaluate calibration, similar to previous work^19^. In causal simulations, we selected 15 causal gene programs, and randomly selected 100 genes connected to these programs as causal genes, and randomly selected 20,000 variants connected to these causal genes and programs as causal variants. In null simulations, we randomly draw *K* causal variants from the total variant set. In causal simulation, to induce functional enrichment^19^, we modified the prior probability that a variant was selected to be causal, setting this to be proportional to Var(*β*_*j*_) = Σ_*C*_*C*(*j*)τ_*C*_ where the estimated enrichment parameters τ_*C*_ were obtained from a meta-analysis of 31 independent traits^31^. For both null and causal simulations, we conducted 500 random replicates. Each replicate consists of different configurations of the causal variants. For the baseline methods, we included the original GWAS (FastGWA/PLINK) and FINDOR^19^. FINDOR uses functional genomics data to identify variants that are more likely to be trait-associated but only incorporate baseline-LD variant-level annotations^31^ such as whether the variant is in an enhancer, coding, or conserved region. For computing independent associations, we merged genome-wide significant variants with LD *r*^2^ > 0.01 within 10MB into one hit and further merged hits within 0.1 centimorgan^53^.

### Experimental details of the subsampling replication analysis

To systematically evaluate small-cohort GWAS on real-world traits, we subsampled a large-cohort trait (*N*=374K) to any small sample size of interest (*N*=1K-10K). We then applied GWAS methods on the small cohort and measured if the associations could be replicated in the large cohort using conventional GWAS at a genome-wide significant threshold. We chose 21 well-powered traits with z-score > 6 for nonzero SNP-heritability and they are selected to have low genetic correlation 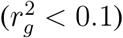 to gauge performance across a wide range of genetic architectures. These traits cover both case-control and continuous phenotypes. For each of the 21 traits, the UK Biobank coding and additional curation step are described in Supplementary Table 5. Five replicates of subsampled cohorts were used to gauge robustness. To estimate the sample size reduction of KGWAS, we used linear interpolation based on the number of discoveries from five sample sizes (1K, 2.5K, 5K, 7.5K, 10K).

### Experimental details of applications to 554 uncommon diseases

We selected ICD10 diseases in UK Biobank (Field ID: 41270) from C01 to R90 with the number of cases > 100. We used Fast-GWA to compute the base GWAS summary statistics. For the GWAS catalog analysis, we mapped the ICD10 code to EFO phenotypes using https://github.com/EBISPOT/EFO-UKB-mappings. We used 5 × 10^−8^ as the threshold of the GWAS catalog associations.

### Experimental details of locus cell type enrichment analysis

Given the variant, we computed the chromatin accessibility signal of 1K base pair +/-region. For each of the 222 cell type tracks^66^, we first conducted a genome-wide z-score normalization per each cell type track and then computed the final z-score by normalizing across cell types at the locus of interest. We then ranked all cell types to obtain the top enriched cell types for the given locus.

### Experimental details of disease network prioritization analysis

For network simulation, we followed the same procedure as in causal simulation and recorded the links used to map from program to genes, genes to genes, and genes to variants as the causal links. We then performed precision-recall analysis given the KGWAS network weights against the causal links. For the real-traits V2G link prioritization analysis, we used 4 diseases with the most manually annotated V2G links in Open Targets Genetics ground truth data. We mapped these 4 diseases to UK Biobank ICD10 codes (E11 for Type II diabetes mellitus, C50 for breast carcinoma, J45 for asthma, and I25 for coronary artery disease). We computed the percentile of KGWAS weights of these ground-truth links in the overall KGWAS variant-to-gene link weights. For the real-trait G2G link prioritization analysis, we used genome-wide perturb-seq data^76^ in K562 hematopoietic cell line and computed ground-truth G2G links if the perturbed gene has statistically significant perturbation effect (FDR<0.05) on the expressed gene. We then applied KGWAS to a K562-related blood trait MCH (Field ID: 30040) and visualized the distribution of KGWAS network weights for the statistically significant links and the rest of the links.

### Experimental details of analysis on molecular mechanisms of Alzheimer’s disease

For the variant interpretation, we obtained the top 3 linked entities with the highest KGWAS link weight for each node type. The ground truth drug target set is curated in Zhang et al.^52^ from Open Targets^88^ where it selected all genes with drug score >0 (clinical trial phase 1 and above). We queried DisGeNet with Alzheimer’s disease (CUI: C0002395) to obtain disease-gene scores. We then aggregated the prioritized/non-prioritized top genes in the first/second hops in the neighborhood to measure enrichment. For the gene program DisGeNet association score, we took the mean of the association scores for the genes in the gene program. We used Cytoscape^100^ to create the network visualization.

### Experimental details of gene prioritization and disease-critical cell population analysis

For gene prioritization replication analysis, we used the same 21 independent traits in the subsampling replication analysis above and applied MAGMA on GWAS/KGWAS summary statistics to generate gene association scores. We then applied the MAGMA on full-cohort GWAS summary statistics and applied a threshold of 0.05 with Bonferroni correction to generate the ground truth gene list. We then evaluated the replication at top *K* = 1000 genes. For drug target genes analysis, the ground truth drug target set is curated in Zhang et al.^52^ from Open Targets^88^ where it selected all genes with drug score >0 (clinical trial phase 1 and above). We then reported the number of replicable top 1000 genes. For disease-critical cell detection analysis, we used scDRS because it is compatible with summary statistics and we input GWAS/KGWAS respectively. Since scDRS requires well-powered summary statistics, we analyzed 93 UK Biobank diseases with >10K cases or *P* <0.05 for non-zero SNP-heritability. We applied on the Tabula Muris Senis scRNA-seq dataset^89^ with 110,824 cells from 120 cell types. We reported the number of detected cells defined as FDR < 0.1. For visualization purposes, we visualized the top 20 cell types with the largest associated cell counts and grouped others.

### User interface KGWAS Explainer

We used R-shiny to design and create the human-KGWAS user interface. For each disease, we reported the KGWAS summary statistics, KGWAS + MAGMA genes, and KGWAS + scDRS cell types. We included a panel of KGWAS diseases critical network where local neighborhoods around genome-wide significant variants are jointly visualized. The edge contains meta-data including edge type and KGWAS weight. An adjustable bar is provided to filter links based on KGWAS network weights. We also included panels to explore KGWAS variants around MAGMA genes and also local network interpretation for each genome-wide significant variant. We included all 726 UK Biobank ICD10 diseases.

## Data availability

The KGWAS knowledge graph and variant/gene/program annotations, summary statistics of GWAS/FINDOR/KGWAS of null, causal simulations, and 21 well-powered independent UK Biobank disease/traits across different sample sizes, summary statistics of GWAS/KGWAS of 554 uncommon diseases are all publicly available at Harvard Dataverse under https://doi.org/10.7910/DVN/C45SO2.

## Code availability

The code to reproduce results, documentation, and usage examples are at https://github.com/snap-stanford/KGWAS. To facilitate the usage of the algorithm, we developed a KGWAS Explainer, a web-based app available at kgwas.stanford.edu to explore KGWAS variants, genes, cell types, and networks.

## Acknowledgements

We thank Jonathan Pritchard, Matthew Aguirre, Yanay Rosen, Yusuf Roohani, Michael Bereket, Rok Sosic, Charlotte Bunne, Hanchen Wang, Minkai Xu, members of Jonathan Pritchard’s lab, and Jure Leskovec’s lab for discussion and providing feedbacks on our manuscript.

This research was conducted using the UK Biobank resource under application no. 79791. K.H. acknowledges support from the Stanford Bio-X Lubert Stryer Interdisciplinary Graduate Fellowship. T.Z. acknowledges support from the Stanford Genetics and Developmental Biology Training Program. J.M.E. acknowledges support from the NHGRI Impact of Genomic Variation on Consortium (UM1HG011972); the Novo Nordisk Foundation Center for Genomic Mechanisms of Disease (NNF21SA0072102); Gordon and Betty Moore and the BASE Research Initiative at the Lucile Packard Children’s Hospital at Stanford University. M.J.Z. acknowledges the support of the Shurl and Kay Curci Foundation. J.L. acknowledges the support of NSF under Nos. OAC-1835598 (CINES), CCF-1918940 (Expeditions), DMS-2327709 (IHBEM), IIS-2403318 (III); Stanford Data Applications Initiative, Wu Tsai Neurosciences Institute, Stanford Institute for Human-Centered AI, Chan Zuckerberg Initiative, Amazon, Genentech, GSK, Hitachi, SAP, and UCB.

## Authors contribution

K.H., C.R., K.B., M.J.Z, J.L. conceived the study. K.H., T.Z., S.K., A.P., C.R., J.M.E., M.J.Z., J.L. performed research, contributed new analytical tools, designed algorithmic frameworks, analyzed data, and wrote the manuscript. K.H., T.Z., and S.K. performed the experiments and designed the software. J.Z., M.J., D.S., C.R., H.R. contributed to code and performed analyses. All authors discussed the results and contributed to the final manuscript.

