## Supplementary Materials for "Small-cohort GWAS discovery with AI over massive functional genomics knowledge graph"

898 # A Supplementary Figures

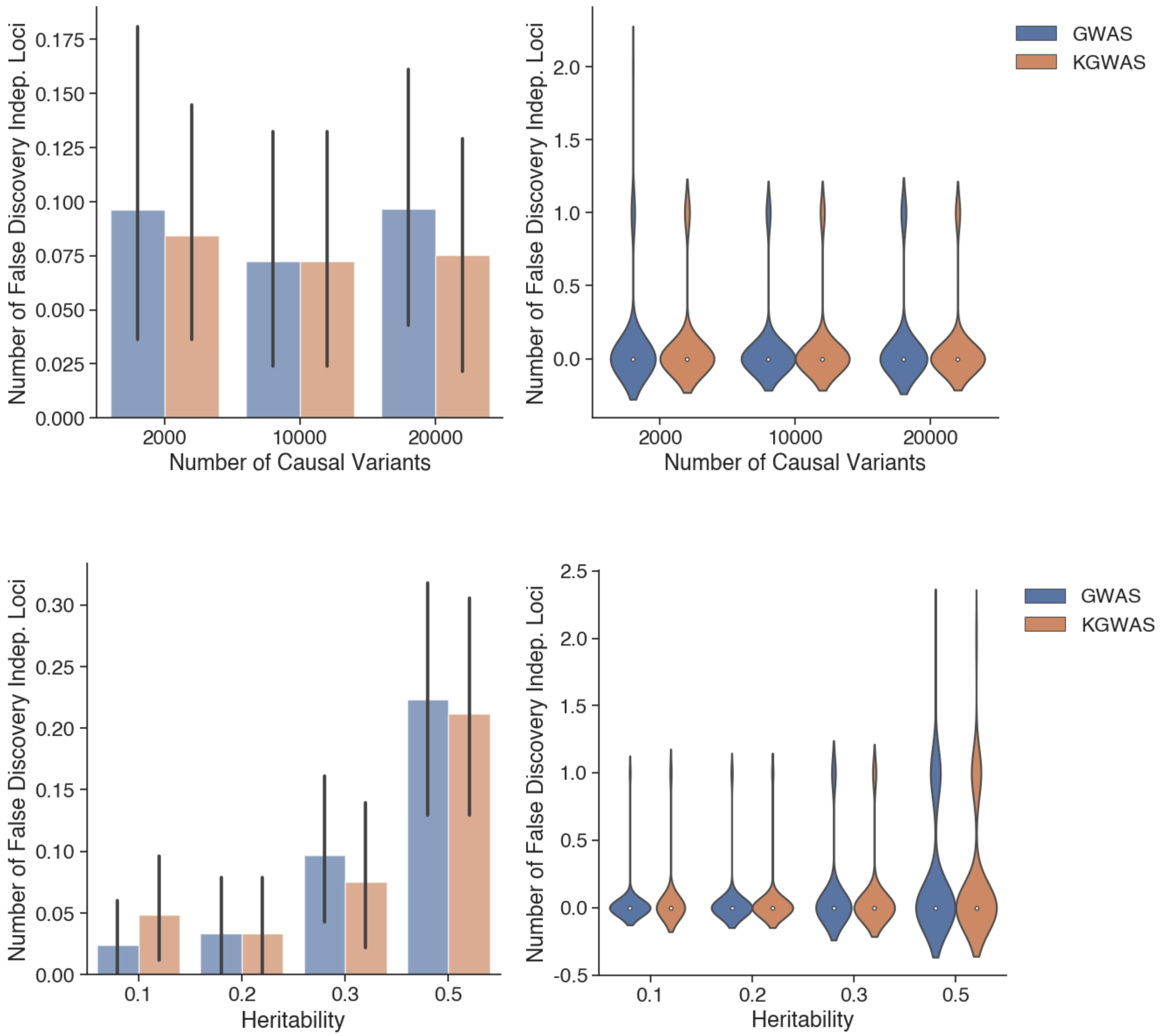


**Supplementary Figure 1: Additional results for null simulations.** Additional configurations of null simulation across a number of variants and heritability show that KGWAS is robust in false discovery control under various genetic architectures. Each configuration is computed with 100 random runs.

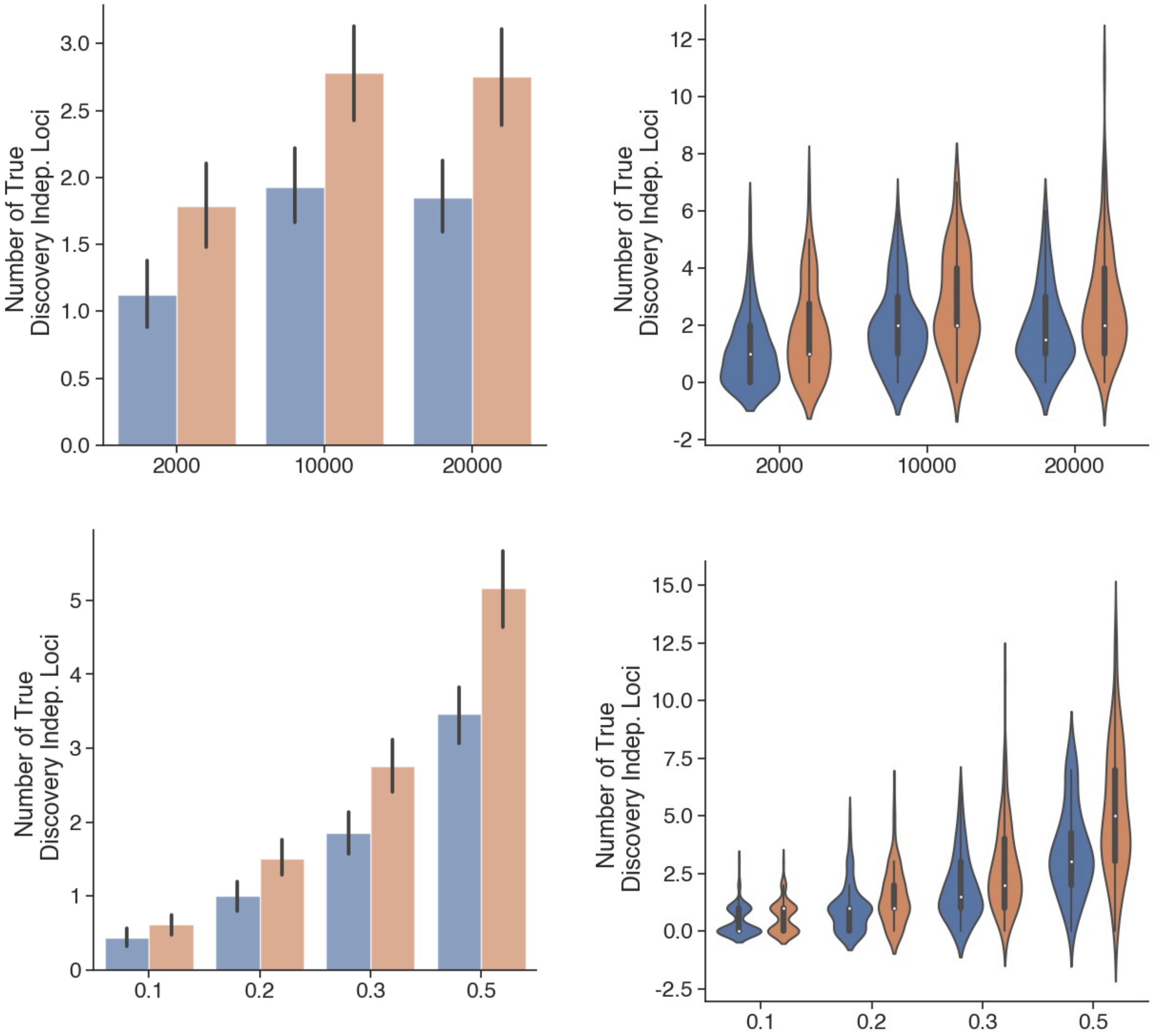


**Supplementary Figure 2: Additional results for causal simulations.** Additional configurations of causal simulation across a number of variants and heritability show that KGWAS is robust in discovering novel loci under various genetic architectures. Each configuration is computed with 100 random runs.

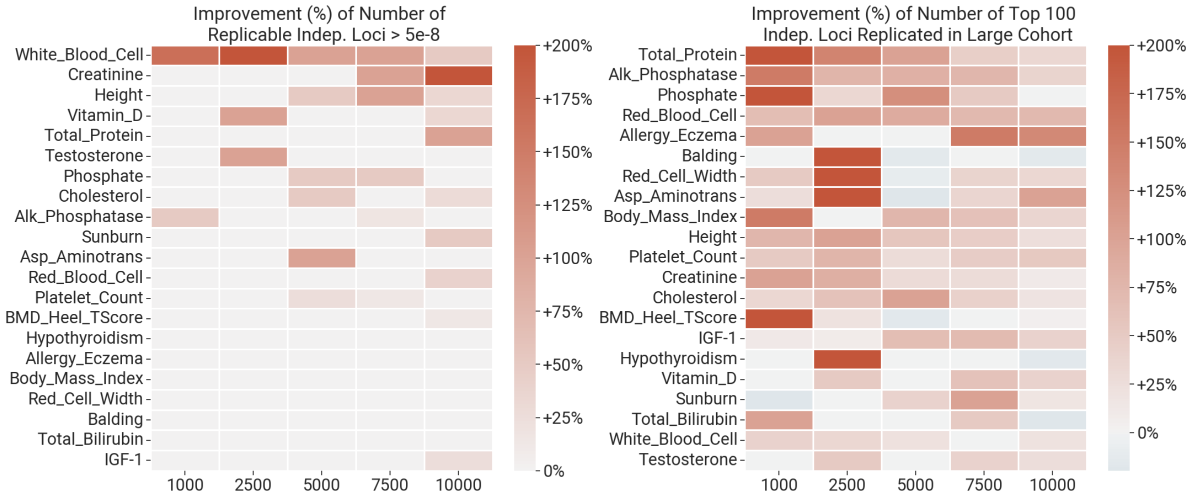


**Supplementary Figure 3: Replication experiment results for individual diseases/traits.** Breakdown of performance on individual traits from the 21 non-redundant traits subsampling analysis. We observe the consistent improvement of KGWAS over GWAS across 21 traits and across all sample sizes for a number of replicable top 100 loci (left panel) and the number of replicable loci at genome-wide significant threshold (right panel).

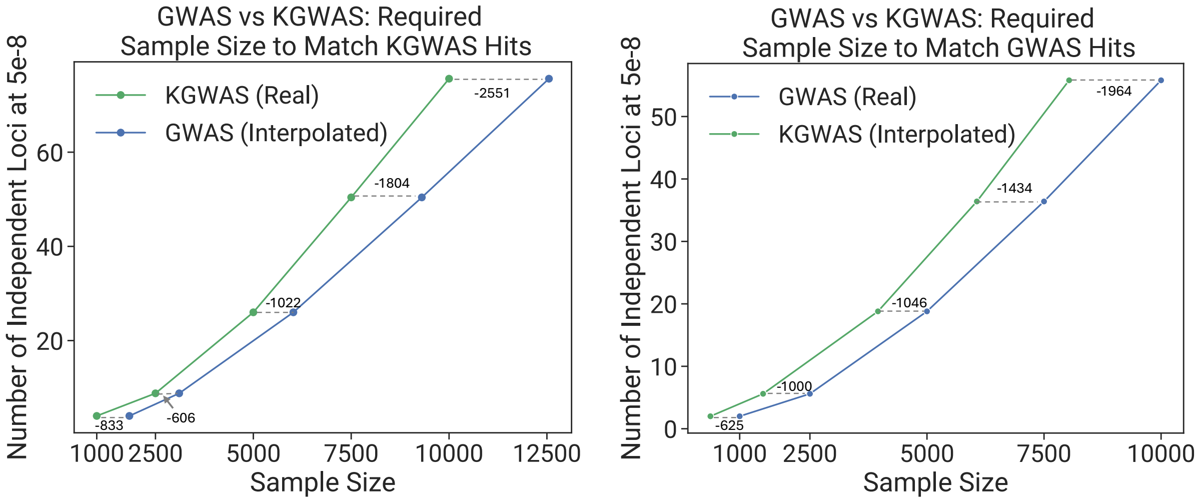


**Supplementary Figure 4: Comparison of data efficiency.** In the 21 independent traits subsampling experiments, for a given GWAS sample size, we estimate the sample size for KGWAS to achieve the same number of discoveries as GWAS. Notably, to achieve the same discoveries as GWAS at 5000 sample size, KGWAS only needs 3954 samples, saving more than 1000 samples. The gain is most prominent in the small cohort.

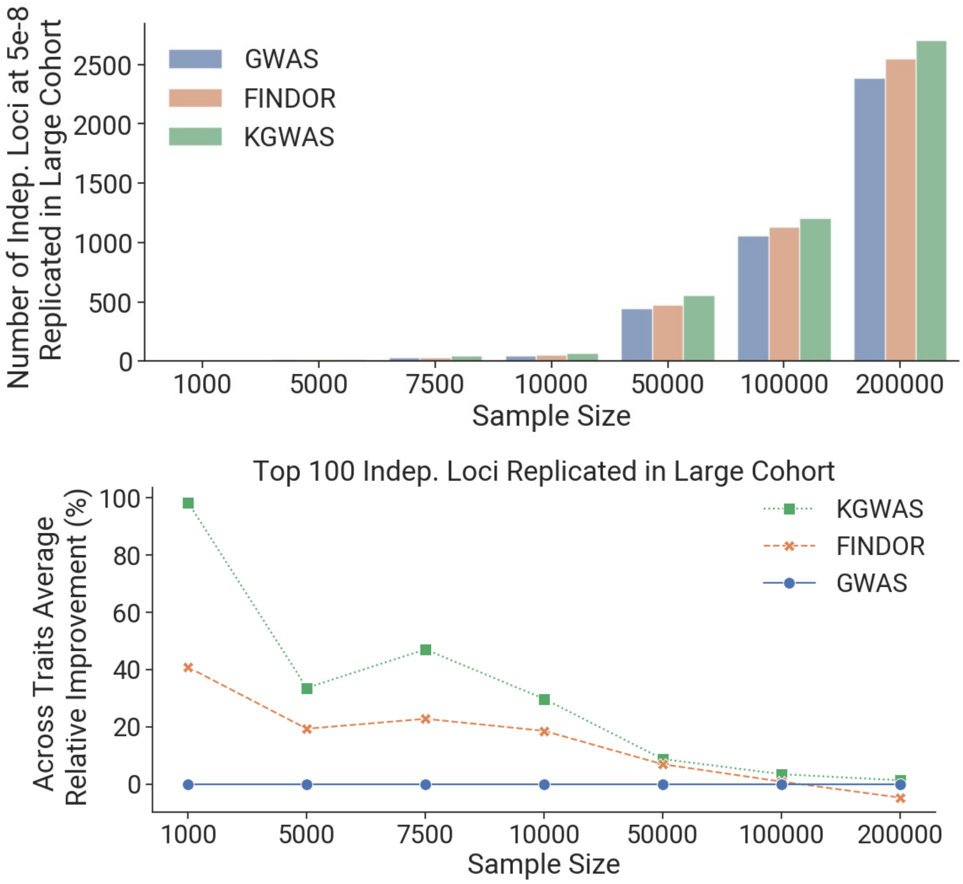


**Supplementary Figure 5: Additional results for replication experiments with larger sample sizes.** For the subsampling analysis, we increase the size of the subsampled cohort to 50K, 100K, and 200K. We observe consistent improvement over GWAS. For the top 100 loci, the results also converge at larger sample sizes. This is because, at larger sample sizes, the top 100 independent loci are easier to detect and consistently form the same set of independent loci, leading to minimal changes in performance.

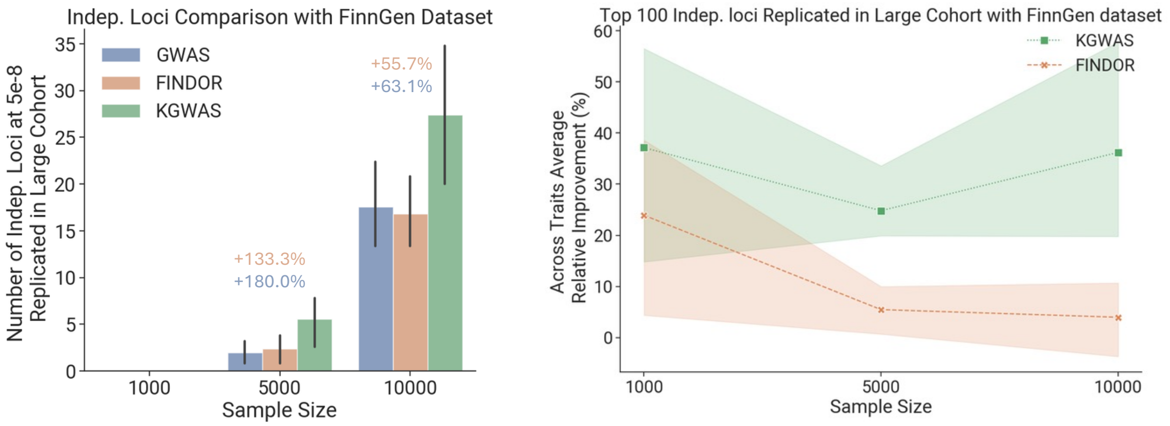


**Supplementary Figure 6: Additional results for replication experiments using the FinnGen cohort.** We replicated the subsampling analysis using the FinnGen cohort. We conduct the same procedure and replicate Figure 2bc. We observe a similar trend in performance. For the trait selection, we first map 4 of 21 non-redundant UKBB traits to FinnGen (Hypothyroidism, strict autoimmune, Dermatitis and eczema, Height, inverse-rank normalized, Body-mass index, inverse-rank normalize) as others do not have direct mapping. For the additional 11 traits, we select based on diversity across five categories: Blood-immune (Autoimmune diseases, Varicose veins), Brain (Depression or dysthymia), Lipid (Type 2 diabetes, definitions combined) and Others (Hypertension, Ischaemic heart disease, wide definition, Asthma/COPD (KELA code 203), Malignant neoplasm of prostate (controls excluding all cancers), Disorders of the thyroid gland, Leiomyoma of uterus, Glaucoma).

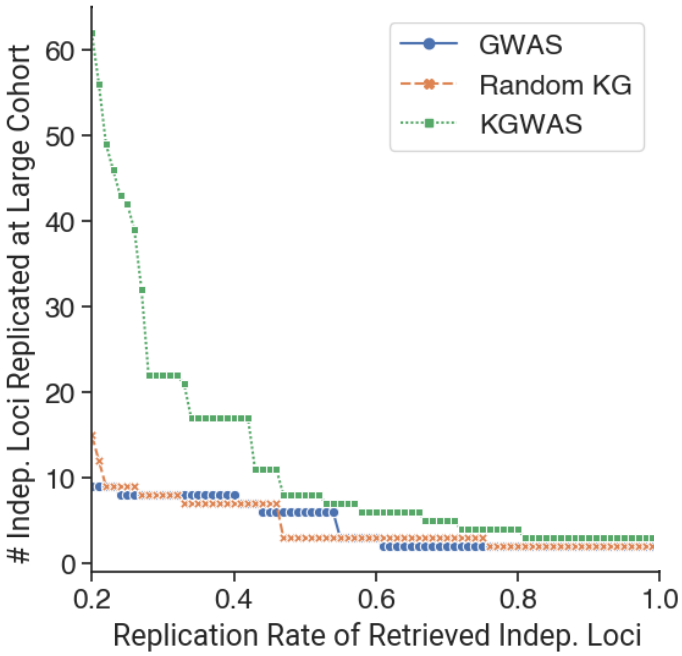


**Supplementary Figure 7: Impact of using a random KG.** We randomize the knowledge graph by randomly permuting edges for every edge type and then performing a subsampling analysis on IGF-1. We observe that by randomizing the KG, the performance degrades to base GWAS, showing that prior knowledge in the KG drives the most performance improvement and the structure in the KG is essential.

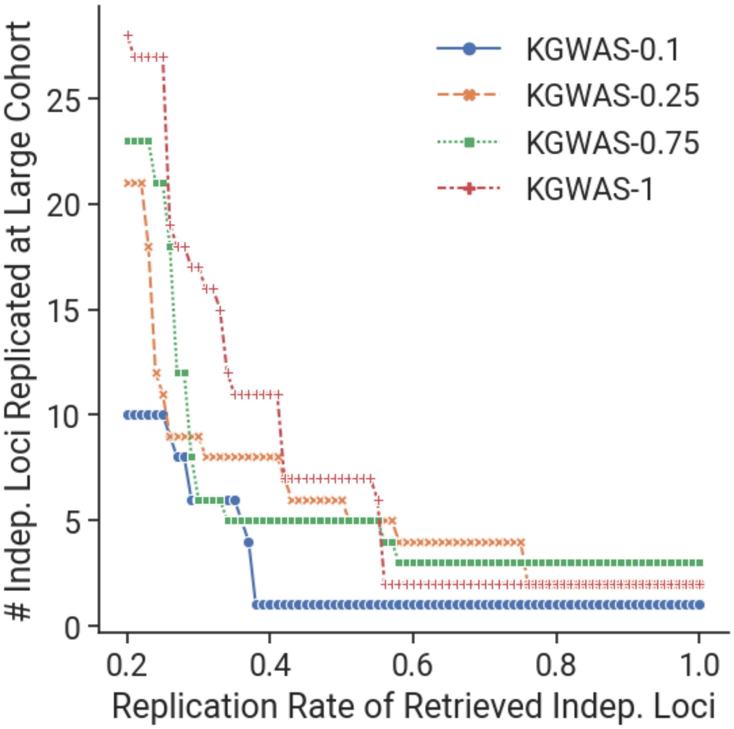


**Supplementary Figure 8: Impact of removing part of the KG.** We randomly remove a fraction of the KG and conduct the subsampling analysis on IGF-1. We show that as the percentage of removal goes up, the performance degrades. This shows the scaling of prior knowledge in the KG and we argue that with more knowledge in the KG, it can potentially further improve the performance.

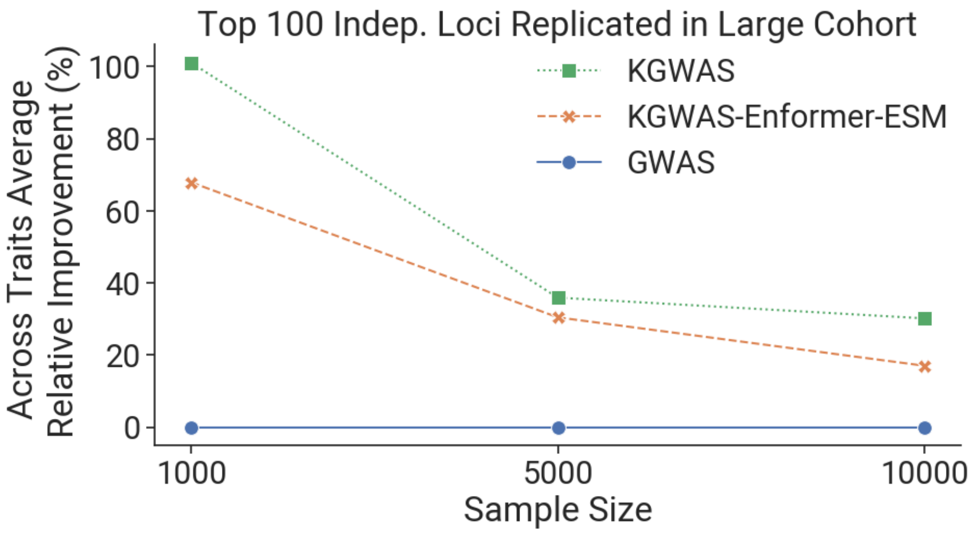


**Supplementary Figure 9: Performance of using alternative node embeddings.** We switched out the initialized embedding of KGWAS from scRNA-seq profiles to ESM embedding and baselineLD features to enformer embedding and then reported the subsampling analysis across three sample sizes. We observe that it has consistent improvement over base GWAS but underperforms compared to KGWAS. We suspect that it is because in human genetics discovery, gene co-expression patterns captured by scRNA-seq is most informative compared to protein structure information in ESM. For the enformer embedding, it is largely capturing functional genomics information, which overlaps with the functional genomics KG. In contrast, a variant-level baselineLD feature provides more orthogonal information.

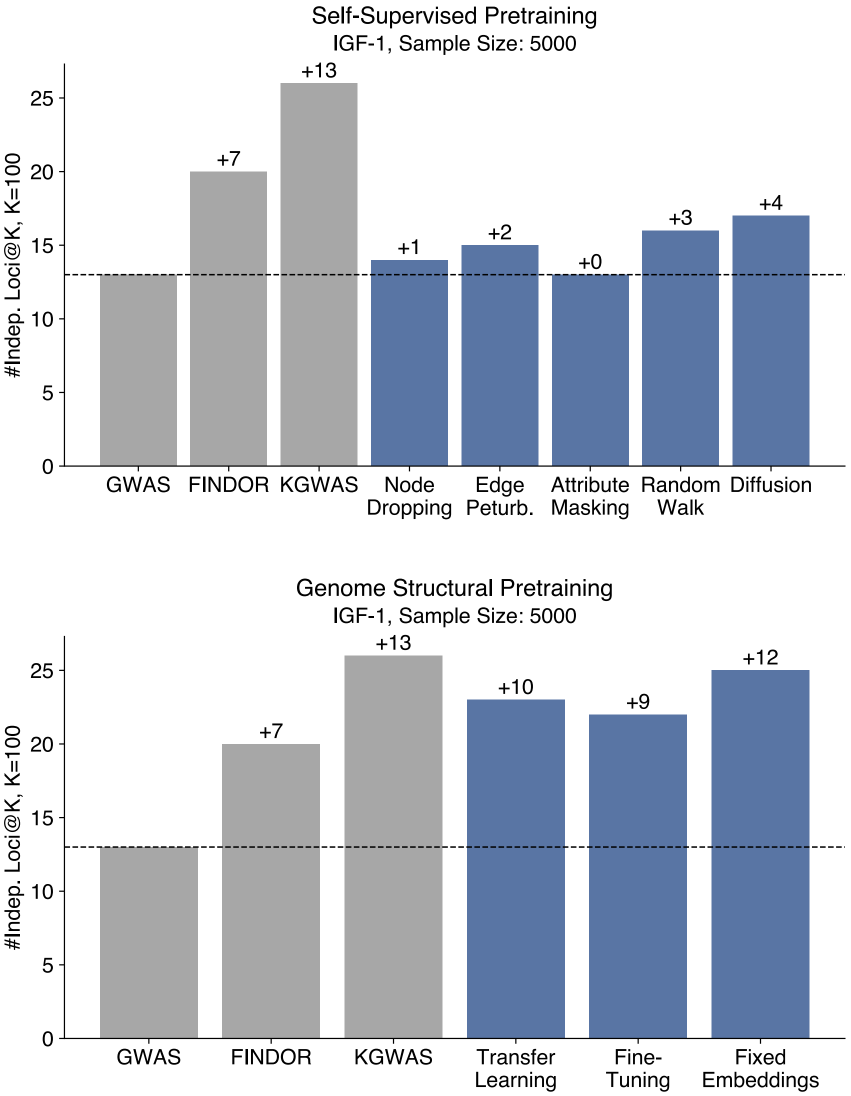


**Supplementary Figure 10: Performance of using alternative pretraining strategies.** KGWAS pre-trained using standard graph self-supervision and a biologically-motivated approach, genome structural pretraining. Pretraining is not seen to improve performance, highlighting the advantage of phenotype-specific training. Graph self-supervision is performed by modifying the minibatch graph at train time either by randomly removing 5% of non-SNP nodes (Node Dropping), removing and adding a random 5% of edges (Edge Perturbation), replacing 10% of node attributes with noise drawn from N(0.5, 0.5) (Attribute Masking), removing nodes not within a length 10 random walk starting from the query SNP (Random Walk), or adding 100 edges between the unconnected node pairs with highest diffusion matrix values (Diffusion). The self-supervision objective is contrastive between the GNN embeddings of the modified and unmodified minibatch graph using the InfoNCE loss with 0.5 temperature. Genome structural pretraining predicts the nucleotide position and chromosome membership of each SNP. The loss is the sum of the MSE of the predicted nucleotide position and cross-entropy of the one-hot-encoded chromosome membership, weighted 1 and 0.1, respectively. Predictions are produced by a 3-layer MLP downstream of the GNN. KGWAS is subsequently trained either with the same learning rate (Transfer Learning), a smaller learning rate of 1e-6 (Fine-Tuning), or fixed GNN embeddings from the pre-trained model (Fixed Embedding).

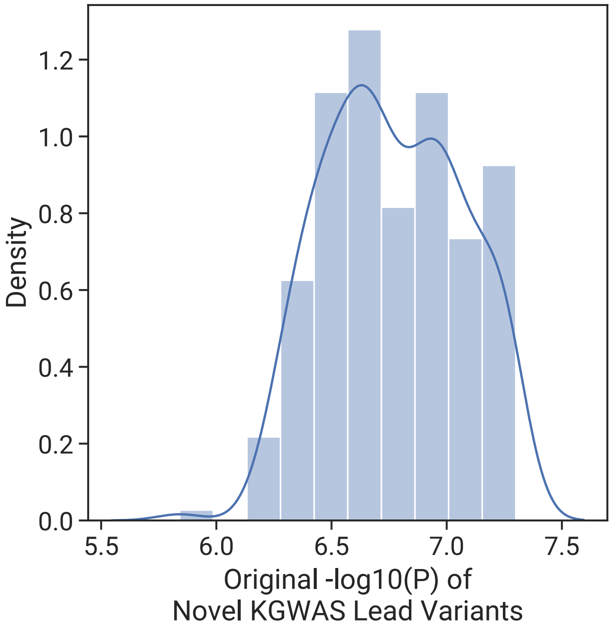


**Supplementary Figure 11: Distribution of the original GWAS p-values for KGWAS-only discoveries.** Original GWAS p-values of KGWAS novel variants range from $1.44 \times 10^{-6}$ to $5.01 \times 10^{-8}$.

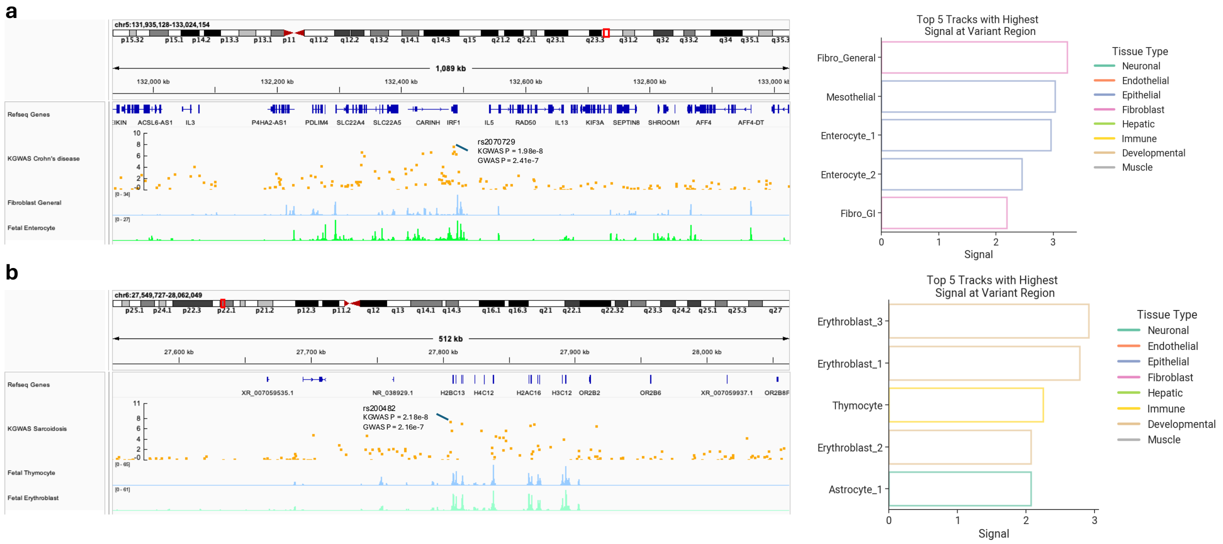


**Supplementary Figure 12: Additional examples of functional evedence for KGWAS-only discoveries. a.** KGWAS novel hit rs2070729 (GWAS $P$=$2.4 \times 10^{-7}$, KGWAS $P$=$2.0 \times 10^{-8}$) for Crohn's disease. Systematic chromatin accessibility analysis suggests it is located in an open region for fibroblasts and enterocytes, both are directly associated with Crohn's disease. **b.** KGWAS novel hit rs200482 (GWAS $P$=$2.2 \times 10^{-7}$, KGWAS $P$=$2.2 \times 10^{-8}$) for Sarcoidosis. Systematic chromatin accessibility analysis suggests it is located in an open region for thymocyte and erythroblast, both are associated with Sarcoidosis.

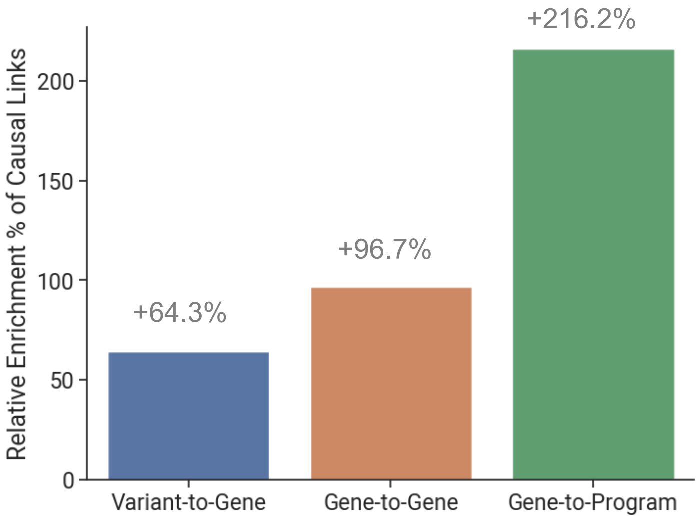


**Supplementary Figure 13: Additional results for KGWAS network link prioritization in causal simulations.** We compute the relative enrichment of KGWAS network weight of the causal links compared to background non-causal links. We observe a consistent improvement of enrichments across all three relation types from variant-to-gene, gene-to-gene, and gene-to-program.

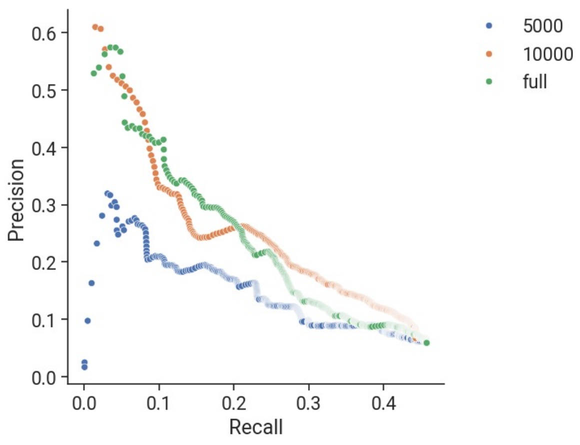


**Supplementary Figure 14: Performance of KGWAS network link prioritization at small sample sizes.** When the sample size is limited, the learned network has a slightly decreased prioritization ability.

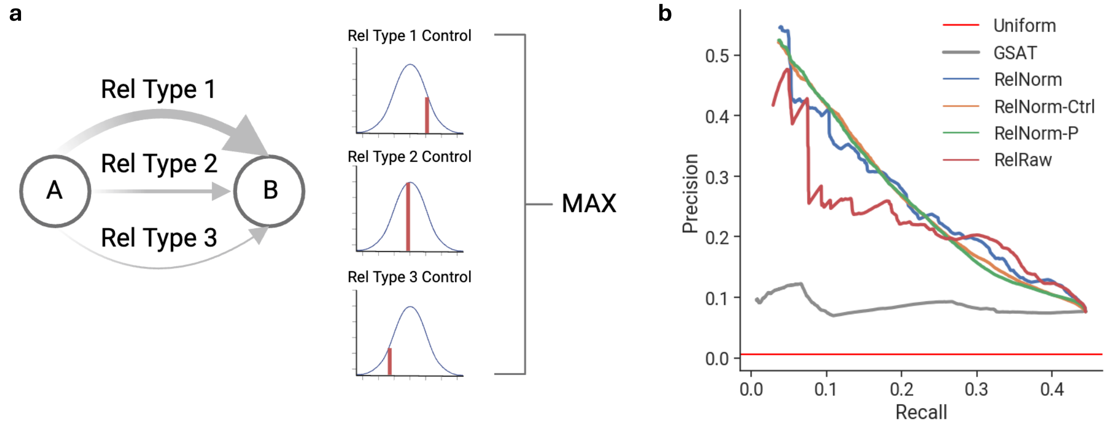


**Supplementary Figure 15: Evaluating alternative strategies for KGWAS network link prioritization. a**. The illustration of how we compute network importance score. For each node A and B, there are multiple relation types, and for each relation type, we obtained a background distribution and computed a z-score for each relation type. We then merge them using the max operator to obtain a score for every pair of nodes. **b**. Benchmarks of variants of KGWAS network importance scores. RelNorm is our current method. RelNorm-ctrl first matches 10 control links for every link and uses that as the control distribution. This produces a more smooth score but it is computationally costly and also similar prioritization performance as the base RelNorm. Thus, we use RelNorm currently. We also strived to use the network control distribution to compute empirical p-values. We found that it is not calibrated but has a similar prioritization performance as RelNorm-ctrl. We also have RelRaw which is without using normalization across relation types. This under-performs the RelNorm, showing the benefit of normalization.

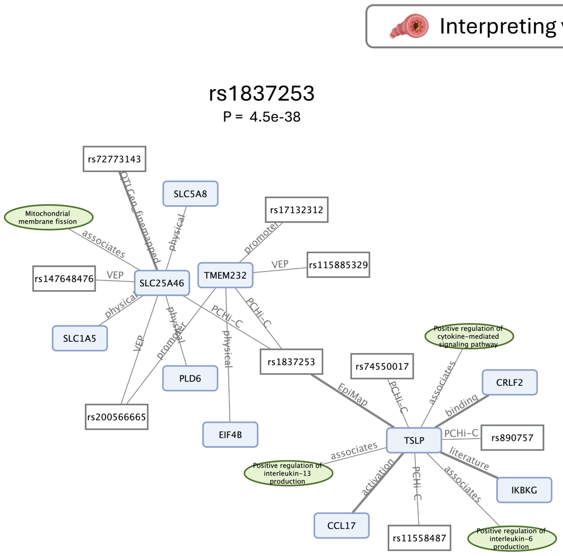


**Supplementary Figure 16:** Example KGWAS network interpretations for asthma. On the left side, the variant rs1837253 has been linked to *TSLP* (via PCHi-C and other functional genomic evidence), a gene central to asthma pathophysiology due to its role in the activation of type 2 innate lymphoid cells and promotion of cytokine-mediated signaling pathways[101]. This connection also implicates downstream genes, such as *CRLF2* and *CCL17*, known for their involvement in immune response regulation, including interleukin production. Furthermore, pathways associated with rs1837253, such as positive regulation of interleukin-6 and -13 production[102], underscore its potential contribution to airway inflammation and hyper-responsiveness in asthma. On the right side, the variant rs2160203 is associated with *IL1RL1* and *IL18RAP* (via PCHi-C and other evidence), both key genes in the immune response. *IL1RL1* plays a crucial role in asthma by mediating type 2 inflammation through its involvement in T-helper 2 (Th2) cytokine production and signaling pathways[103]. Additionally, *IL18RAP* links this variant to the interleukin-18-mediated signaling pathway, further emphasizing its role in inflammation and airway hyper-responsiveness. Pathways like the positive regulation of NF-$\kappa$B transcription factor activity and type II interferon production highlight its potential contributions to the chronic inflammatory environment characteristic of asthma.

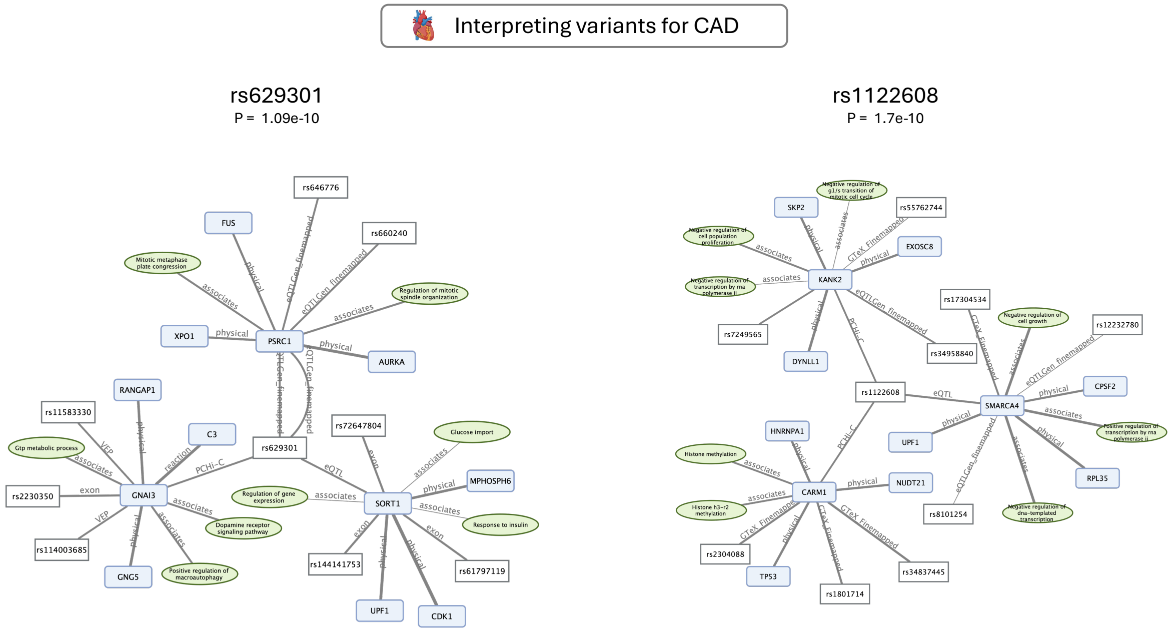


**Supplementary Figure 17:** Example KGWAS network interpretations for coronary artery disease. On the left side, the variant rs629301 is linked to *PSRC1* and *SORT1* (via PCHi-C and additional genomic evidence), genes implicated in lipid metabolism and coronary artery disease (CAD). *PSRC1* is associated with pathways regulating mitotic spindle organization and progression, suggesting its role in cell cycle regulation and vascular health[104]. *SORT1*, on the other hand, is involved in glucose import and insulin response, connecting this variant to metabolic pathways critical in CAD development[105]. Additional associations with genes like *GNAI3*, which participates in dopamine receptor signaling and macroautophagy regulation, highlight potential mechanisms by which this variant could contribute to CAD risk through metabolic and inflammatory dysregulation. On the right side, the variant rs1122608 is linked to *KANK2* and *SMARCA4* (via PCHi-C and eQTL evidence), both of which are involved in processes critical to coronary artery disease (CAD). *KANK2* is associated with the regulation of cell population proliferation and the G1/S transition of the mitotic cell cycle, suggesting its potential role in vascular smooth muscle cell growth and plaque stability[106]. *SMARCA4*, a regulator of chromatin remodeling, is connected to pathways such as transcription regulation and cell growth inhibition, underscoring its role in maintaining cellular homeostasis and preventing vascular remodeling[107]. Additionally, connections to *CARM1*, involved in histone H3 methylation, highlight potential epigenetic mechanisms influencing CAD susceptibility[108]. These findings suggest that rs1122608 impacts CAD risk through a combination of transcriptional regulation, epigenetic modification, and cell cycle control.

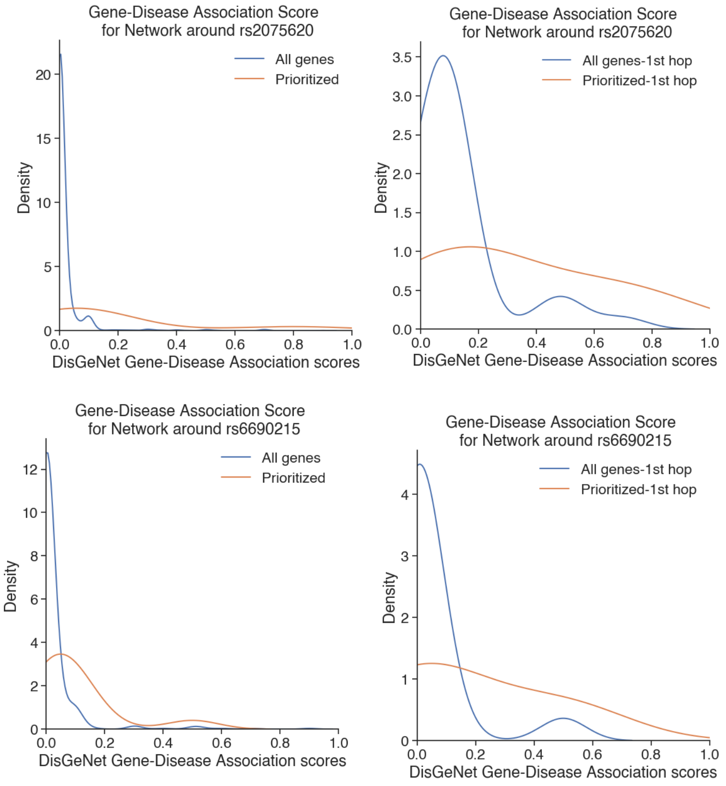


**Supplementary Figure 18: Enrichment of DisGeNet scores for KGWAS-prioritized genes.** The prioritized network interpretation has enriched genes that are associated with the disease. For rs2075620, the average GDA score for all 2-hop neighborhoods is 0.018 and for the KGWAS prioritized 2-hop neighborhood is 0.182, a 10.1 fold enrichment. The average GDA score for a 1-hop neighborhood is 0.128 and for the KGWAS prioritized 1-hop neighborhood is 0.323, a 2.5-fold enrichment. Similarly, for rs6690215, the average GDA score for all 2-hop neighborhoods is 0.025 and for the KGWAS prioritized 2-hop neighborhood is 0.095, a 3.8-fold enrichment. The average GDA score for a 1-hop neighborhood is 0.048 and for the KGWAS prioritized 1-hop neighborhood is 0.180, a 3.75 fold enrichment.

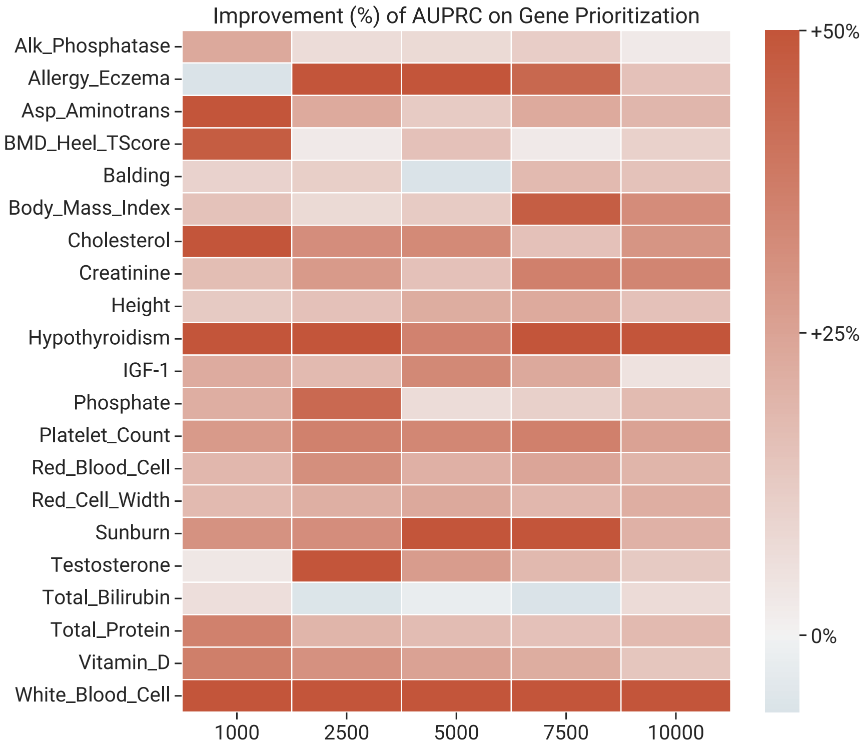


**Supplementary Figure 19: Additional results for disease gene prioritization.** We report the trait-specific performance for gene-level systematic subsampling replication study on AUPRC and top 1000 genes replication. KGWAS consistently outperforms GWAS across traits and sample sizes.

### B Supplementary Tables

| Node types | Number of nodes |
|---|---|
| Variant | 784256 |
| Gene | 23737 |
| CellularComponent | 4184 |
| MolecularFunction | 11153 |
| BiologicalProcess | 28748 |

**Supplementary Table 1: KG node statistics.** We report the type of nodes and the number of nodes for each type. "CellularComponent", "MolecularFunction", and "BiologicalProcess" are gene program nodes in the knowledge graph.

See Supplementary Excel file.

**Supplementary Table 2: KG relation statistics.** We report the type of edges and the number of edges for each type in the knowledge graph.

| Method | # False Pos. |
|---|---|
| FINDOR | 0.018±0.1329 |
| GWAS | 0.016±0.1254 |
| KGWAS | 0.018±0.1329 |

**Supplementary Table 3: Null simulation results.** We report the numerical experimental results for the null simulation.

| Method | # of significant indep. loci |
|---|---|
| FINDOR | 2.51±1.5715 |
| GWAS | 1.85±1.4097 |
| KGWAS | 2.75±1.8834 |

**Supplementary Table 4: Causal simulation results.** We report the numerical experimental results for the causal simulation.

| Trait Name | Trait_Identifier | UKBB Column Codes | Additional information on coding | Prevalence | h2g | sd | Z |
|---|---|---|---|---|---|---|---|
| Eczema | UKB_460K.disease_ALLERGY_ECZEMA_DIAGNOSED | 6152-0.0∼6152-0.4 | Hayfever, allergic rhinitis or eczema diagnosed by doctor | 0.2312 | 0.0847 | 0.0058 | 14.60344828 |
| Heel T Score | UKB_460K.bmd_HEEL_TSCOREz | 4106-0.0+4125-0.0 | - | | 0.3549 | 0.0173 | 20.51445087 |
| Hypothyroidism | UKB_460K.disease_HYPOTHYROIDISM_SELF_REP | 20002-0.0∼20002-0.28 | 1226 | 0.04824 | 0.0546 | 0.0036 | 15.16666667 |
| Balding Type I | UKB_460K.body_BALDING1 | 2395-0.0 | balding pattern 1 vs. other | 0.32 | 0.2234 | 0.0152 | 14.69736842 |
| Sunburn Occasion | UKB_460K.pigment_SUNBURN | 1737-0.0 | any sunburn occasion or not | 0.46 | 0.0731 | 0.0076 | 9.618421053 |
| AlkalinePhosphatase | UKB_460K.biochemistry_AlkalinePhosphatase | 30610-0.0+30897-0.0 | remove ID with value outside mean +/-4sd | | 0.2348 | 0.016 | 14.675 |
| AspartateAminotransferase | UKB_460K.biochemistry_AspartateAminotransferase | 30650-0.0+30897-0.0 | remove ID with value outside mean +/-4sd | | 0.1135 | 0.0062 | 18.30645161 |
| Cholesterol | UKB_460K.biochemistry_Cholesterol | 30690-0.0+30897-0.0 | remove ID with value outside mean +/-4sd | | 0.1307 | 0.0112 | 11.66964286 |
| Creatinine | UKB_460K.biochemistry_Creatinine | 30700-0.0+30897-0.0 | remove ID with value outside mean +/-4sd | | 0.2213 | 0.0101 | 21.91089109 |
| IGF1 | UKB_460K.biochemistry_IGF1 | 30770-0.0+30897-0.0 | remove ID with value outside mean +/-4sd | | 0.2877 | 0.0154 | 18.68181818 |
| Phosphate | UKB_460K.biochemistry_Phosphate | 30810-0.0+30897-0.0 | remove ID with value outside mean +/-4sd | | 0.1292 | 0.0091 | 14.1978022 |
| TestosteroneMale | UKB_460K.biochemistry_Testosterone_Male | 30850-0.0+30897-0.0 | remove ID with value outside mean +/-4sd | | 0.184 | 0.0111 | 16.57657658 |
| TotalBilirubin | UKB_460K.biochemistry_TotalBilirubin | 30840-0.0+30897-0.0 | remove ID with value outside mean +/-4sd | | 0.0841 | 0.0076 | 11.06578947 |
| TotalProtein | UKB_460K.biochemistry_TotalProtein | 30860-0.0+30897-0.0 | remove ID with value outside mean +/-4sd | | 0.1811 | 0.0075 | 24.14666667 |
| VitaminD | UKB_460K.biochemistry_VitaminD | 30890-0.0+30897-0.0 | remove ID with value outside mean +/-4sd | | 0.086 | 0.0069 | 12.46376812 |
| BMI (UK Biobank) | UKB_460K.body_BMIz | 21001-0.0 | - | | 0.2779 | 0.0067 | 41.47761194 |
| Height (UK Biobank) | UKB_460K.body_HEIGHTz | 50-0.0 | - | | 0.674 | 0.0261 | 25.82375479 |
| Platelet Count | UKB_460K.blood_PLATELET_COUNT | 30080-0.0 | - | | 0.3505 | 0.0165 | 21.24242424 |
| Red Blood Cell Distribution Width | UKB_460K.blood_RBC_DISTRIB_WIDTH | 30070-0.0 | - | | 0.2172 | 0.0122 | 17.80327869 |
| Red Blood Cell Count | UKB_460K.blood_RED_COUNT | 30010-0.0 | - | | 0.2613 | 0.012 | 21.775 |
| White Blood Cell Count | UKB_460K.blood_WHITE_COUNT | 30000-0.0 | - | | 0.2273 | 0.0083 | 27.38554217 |

**Supplementary Table 5: 21 non-redundant traits used in subsampling analysis.** We report the descriptions, curations, prevalence, heritability of the 21 non-redundant traits used in the subsampling replication experiment.

See Supplementary Excel file.

**Supplementary Table 6: Subsampling analysis results at genome-wide significance threshold.** We report the numerical experimental results on the number of replicable independent loci that pass the genome-wide significant threshold for the subsampling replication analysis for each trait.

See Supplementary Excel file.

**Supplementary Table 7: Subsampling analysis raw results at top 100 independent loci.** We report the numerical experimental results on the number of replicable independent loci in the top 100 loci for the subsampling replication analysis for each trait.

See Supplementary Excel file.

**Supplementary Table 8: Results for 554 uncommon diseases in UK BioBank.** We report the list of 554 uncommon diseases, the number of cases for each disease, and the number of genome-wide significant discoveries for KGWAS and GWAS.

See Supplementary Excel file.

**Supplementary Table 9: MG CATLAS cell type enrichment statistics.** We report the cell type signal enrichment score around the KGWAS MG locus for 222 cell types in the CATLAS data.

See Supplementary Excel file.

**Supplementary Table 10: Causal network prioritization simulation results.** We report the numerical results (precision and recall) of the causal network prioritization simulation experiment.

See Supplementary Excel file.

**Supplementary Table 11: Causal link variant-to-gene with OpenTarget results.** We report the numerical results of the variant-to-gene experiment. We report the percentiles for each ground truth V2G link across the four diseases.

See Supplementary Excel file.

**Supplementary Table 12: Causal link gene-to-gene with perturb-seq results.** We report the numerical results of the gene-to-gene validation experiment. We report the raw link weights for the G2G link with perturb-seq evidence and rest of the G2G links respectively.

See Supplementary Excel file.

**Supplementary Table 13: AD annotation result for rs2075620.** We report the numerical results for the DisGeNet gene-disease association scores for rs2075620 at 1 and 2 hops of the neighborhood for baseline and KGWAS prioritized genes and programs.

See Supplementary Excel file.

**Supplementary Table 14: AD annotation result for rs6690215.** We report the numerical results for the DisGeNet gene-disease association scores for rs6690215 at 1 and 2 hops of the neighborhood for baseline and KGWAS prioritized genes and programs.

| Trait (medium) | GWAS | KGWAS | Relative Improvement (%) |
|---|---|---|---|
| BMD_Heel_TScore | 13.0 | 14.0 | 7.6923076923076900 |
| Vitamin_D | 13.0 | 14.0 | 7.6923076923076900 |
| Phosphate | 13.0 | 15.0 | 15.384615384615400 |
| Allergy_Eczema | 12.0 | 17.0 | 41.66666666666670 |
| Asp_Aminotrans | 17.0 | 17.0 | 0.0 |
| Balding | 19.0 | 19.0 | 0.0 |
| Hypothyroidism | 7.0 | 21.0 | 200.0 |
| White_Blood_Cell | 9.0 | 21.0 | 133.33333333333300 |
| Total_Bilirubin | 24.0 | 24.0 | 0.0 |
| Body_Mass_Index | 20.0 | 26.0 | 30.0 |
| Sunburn | 21.0 | 26.0 | 23.809523809523800 |
| Testosterone | 28.0 | 29.0 | 3.571428571428570 |
| Cholesterol | 42.0 | 48.0 | 14.285714285714300 |
| IGF-1 | 46.0 | 49.0 | 6.521739130434780 |
| Red_Cell_Width | 43.0 | 54.0 | 25.581395348837200 |
| Creatinine | 46.0 | 57.0 | 23.91304347826090 |
| Alk_Phosphatase | 54.0 | 60.0 | 11.11111111111110 |
| Red_Blood_Cell | 46.0 | 60.0 | 30.434782608695700 |
| Platelet_Count | 68.0 | 75.0 | 10.294117647058800 |
| Height | 70.0 | 84.0 | 20.0 |
| Total_Protein | 76.0 | 91.0 | 19.736842105263200 |

**Supplementary Table 15: Gene replication results.** We report the numerical results for the gene prioritization replication experiments across 21 independent traits for both GWAS and KGWAS.

| Mapped ICD10 code | Top 1000-GWAS | Top 1000-KGWAS | Trait |
|---|---|---|---|
| K50 | 8.0 | 12.0 | CD |
| K90 | 1.0 | 1.0 | Celiac |
| K51 | 10.0 | 11.0 | UC |
| M32 | 5.0 | 6.0 | SLE |
| G35 | 11.0 | 12.0 | MS |
| L30 | 3.0 | 1.0 | Eczema |
| J45 | 15.0 | 12.0 | ASM |
| G30 | 9.0 | 17.0 | AD |
| F31 | 12.0 | 17.0 | BP |
| G47 | 5.0 | 8.0 | Insomnia |
| F32 | 9.0 | 10.0 | MDD |
| F20 | 15.0 | 18.0 | SCZ |
| I48 | 14.0 | 16.0 | AF |
| I25 | 14.0 | 16.0 | CAD |
| I10 | 20.0 | 20.0 | HTN |

**Supplementary Table 16: Gene drug target results.** We report the numerical results for the recall rate between the prioritized GWAS/KGWAS genes and known external drug targets.

See Supplementary Excel file.

**Supplementary Table 17: List of diseases in the disease-critical cell population detection analysis.**

See Supplementary Excel file.

**Supplementary Table 18: Disease-critical cell population result.** We report the numerical result for the number of detected disease-critical cells in all cell types in Tabula Muris when integrating GWAS/KGWAS with scDRS.

| **Task** | **List** | **Rationale** |
|---|---|---|
| Figure 2b-d, 3d, Systematic Replication | 21 traits in UK Biobank | Well-powered in full sample size, independent with each other, $z$-score $> 6$ for nonzero SNP-heritability, $r_g^2 < 0.1$ |
| Figure 3a-b KGWAS applications to small cohort | 554 UK Biobank diseases | KGWAS works the best in small-cohort GWAS and thus we include all diseases in UK-Biobank with $< 5K$ cases |
| Figure 4b-c network interpreter real traits experiment | 5 UK Biobank diseases & traits | Match available traits in the ground truth data |
| Figure 4d-e network interpretations | 3 publicly available large-cohort diseases | Network interpreter works the best when well-powered; thus, we solicit large-cohort GWAS summary statistics file |
| Figure 5b disease target prioritization | 15 UK Biobank diseases | Traits with ground truth disease target set overlap |
| Figure 5c cell-type association detection | 93 UK Biobank diseases | scDRS requires well-powered diseases and thus we use all diseases in UK Biobank with 10K cases or nominal heritability ($P < 0.05$) |
| Figure 5e KGWAS UI interface | 726 UK Biobank diseases | Apply to all diseases in UK Biobank for the user interface |
| Supplementary Figure 10 FinnGen systematic replication | 15 FinnGen traits | For replication of the subsampling framework in another cohort |

**Supplementary Table 19: Summary of traits.** We report the list of diseases/traits used for each analysis in the main figures.
